# Identifying recent cholera infections using a multiplex bead serological assay

**DOI:** 10.1101/2022.06.27.22276845

**Authors:** Forrest K. Jones, Taufiqur R. Bhuiyan, Rachel Mills, Ashraful I Khan, Damien Slater, Kian Robert Hutt Vater, Fahima Chowdhury, Meagan Kelly, Peng Xu, Pavol Kováč, Rajib Biswas, Mohammad Kamruzzaman, Edward T. Ryan, Stephen B. Calderwood, Regina C. LaRocque, Justin Lessler, Richelle C. Charles, Daniel T. Leung, Firdausi Qadri, Jason B. Harris, Andrew S. Azman

**Affiliations:** Department of Epidemiology, Johns Hopkins Bloomberg School of Public Health, Baltimore, USA; Infectious Diseases Division, International Centre for Diarrhoeal Disease Research, Bangladesh (icddr,b), Dhaka, Bangladesh; Division of Infectious Diseases, Massachusetts General Hospital, Boston, USA; NIDDK, LBC, National Institutes of Health, Bethesda, MD, USA; Department of Medicine, Harvard Medical School, Boston, USA; Department of Immunology and Infectious Diseases, Harvard T.H. Chan School of Public Health, Boston, USA; Department of Epidemiology, University of North Carolina Gillings School of Global Public Health, Chapel Hill, USA; University of North Carolina Population Center, University of North Carolina Gillings School of Global Public Health, Chapel Hill, USA; Division of Infectious Diseases, University of Utah School of Medicine, Salt Lake City, USA; Division of Microbiology and Immunology, University of Utah School of Medicine, Salt Lake City, USA; Department of Pediatrics, Harvard Medical School, Boston, USA; Institute of Global Health, University of Geneva, Geneva, Switzerland

**Keywords:** serosurveillance, multiplex bead assay, seroincidence, *Vibrio cholerae*

## Abstract

**Background:** Estimates of incidence based on medically attended cholera can be severely biased. *Vibrio cholerae* O1 leaves a lasting antibody signal and recent advances show that these can be used to estimate infection incidence rates from cross-sectional serologic data. Current laboratory methods are resource intensive and challenging to standardize across laboratories. A multiplex bead assay (MBA) could efficiently expand the breadth of measured antibody responses and improve seroincidence accuracy.

**Methods:** We tested 305 serum samples from confirmed cholera cases (4-1083d post-infection) and uninfected contacts in Bangladesh using an MBA (IgG/IgA/IgM for 7 *Vibrio cholerae* O1-specific antigens) as well as traditional vibriocidal and enzyme-linked immunosorbent assays (2 antigens, IgG and IgA).

**Results:** While post-infection vibriocidal responses were larger than other markers, several MBA-measured antibodies demonstrated robust responses with similar half-lives. Random forest models combining all MBA antibody measures allowed for accurate identification of recent cholera infections (e.g. past 200 days) including a cross-validated AUC (cvAUC_200_) of 92% with simpler 3 IgG antibody models having similar accuracy. Across infection windows between 45- and 300-days, accuracy of models trained on MBA measurements were non-inferior to models based on traditional assays.

**Conclusions:** Our results illustrate a scalable cholera serosurveillance tool that can be incorporated into multi-pathogen serosurveillance platforms.

## Introduction

Cholera remains a global public health threat with an estimated 95,000 deaths per year, especially in areas without access to safe water and adequate sanitation [1]. Seventh pandemic strains of *Vibrio cholerae (*toxigenic serogroup O1 El Tor biotype) are responsible for most cholera cases, with endemic transmission in parts of Africa and Asia as well as large outbreaks in conflict zones, humanitarian crises and post-disaster settings [2–4]. Several countries plan to achieve large reductions in cholera cases and deaths over the next decade using a multisectoral approach that includes administration of oral cholera vaccines and investment in water and sanitation infrastructure [5]. A clear understanding of the magnitude of pandemic *V. cholerae* transmission at the sub-national level is essential for targeting and monitoring of global progress towards ending cholera.

Cholera surveillance typically consists of clinic-based syndromic surveillance for acute watery diarrhea with infrequent laboratory confirmation [6]. When laboratory confirmation is performed, often less than half of suspected cholera cases have detectable *V. cholerae* by culture, though this varies considerably by setting [7–9]. Since most infections with *V. cholerae* lead to mild or no symptoms, clinical surveillance detects only a small fraction of infections [10,11]. Clinical surveillance systems are also subject to biases related to individual healthcare access and design of the surveillance system (e.g. sentinel sites) [12,13]. As a result, clinical surveillance alone provides a skewed understanding of disease burden and transmission of pandemic *V. cholerae*.

Serosurveillance has been a useful complement to clinical surveillance for a variety of pathogens and there is growing interest in its use for monitoring cholera incidence [14,15]. Despite variability in clinical outcomes, infection with cholera, regardless of symptoms, typically leads to a robust measurable immune response. This includes a rise and eventual decay in serum-circulating antibodies against multiple epitopes [10,16]. As a result, cross-sectional measurements of circulating antibodies can provide insights into the incidence and timing of past infections. The two most common methodologies used to measure antibodies generated in response to *V. cholerae* infection are the vibriocidal assay and enzyme linked immunosorbent assay (ELISA). Previous studies have shown that vibriocidal titers rise quickly after infection and then decay toward pre-infection levels after 1 year [16,17]. However, the vibriocidal method is a functional assay that requires culturing *V. cholerae* over several hours (thus requiring Biosafety Level 2 facility) and is challenging to standardize across laboratories [18]. Although ELISAs targeting immunoglobulin G (IgG) and IgA antibodies binding to known antigens are easier to implement, these assays are less predictive of recent infection than the vibriocidal assay [19]. Previous work illustrates that combining vibriocidal titers with ELISA antibody measurements in statistical models can identify recently infected individuals for estimating cholera seroincidence (i.e., the incidence of meaningful immunologic exposures to *V. cholerae* O1 over a specific time period) [19].

Over the past decade, advances in high-throughput serological multiplex bead assays (MBAs) have enabled their use to study the burden, risk, and dynamics of a variety of pathogens [20–23]. These assays only require a small volume of serum (e.g., 1 μL to measure multiple antigens [when performed in duplicate] as compared to 12.5 μL for the vibriocidal assay), potentially are more sensitive [24], and could be easier to standardize [25]. Additionally, they allow for the characterization of large numbers of antigens simultaneously, improving the efficiency and cost of the assay as compared to running multiple ELISAs [26]. This also facilitates broad exploration of novel antigens that may correlate with previous exposure or immunity. If measuring multiple antibodies to *V. cholerae* antigens is as predictive of recent infection as the vibriocidal assay, serosurveillance would be feasible in many more settings. However, the use of cholera antigens in an MBA to predict recent infection has not been previously assessed.

Here, we characterize post-infection antibody dynamics to seven cholera antigens up to three years post-infection in a cohort of confirmed medically-attended *V. cholerae* O1 infections. We use these serological data to train statistical models to identify recently infected individuals. We then compare performance of models based on this assay to those based on traditional antibody measurements and suggest a reduced panel of antigens to be used in MBA arrays for future cholera serosurveillance efforts.

## Materials and Methods

### Study Population

As described previously, consenting patients hospitalized at the icddr,b (formerly known as International Centre for Diarrhoeal Disease Research, Bangladesh) Dhaka hospital with culture-confirmed *V. cholerae* O1 infection were enrolled between 2006 and 2018 [27,28]. These cases were followed up to 3 years post enrollment with blood samples collected periodically. We approximated the number of days between infection and sample collection by taking the difference between the enrollment date and sample collection date, then adding the number of hours of symptomatic diarrhea prior to enrollment (range: 3-60 hours), then adding 1.4 days for the incubation period [29], and finally rounding to a whole number of days. Household contacts of confirmed cases (defined as individuals who shared a cooking pot with the index case for three or more days preceding the cholera episode in the index case) were also enrolled with blood and stool samples collected at approximately 2-, 7-, and 30-days post enrollment of the initial cases. We limited our selection of household contacts to those that had no evidence of cultured *V. cholerae* from stool samples during follow-up. Data on non-immunological variables of age, sex, and blood type were available for all participants. We selected a set of 305 samples from 51 individuals to test and compare to previously measured antibody responses (Supplementary Methods, Figure S1, Figure S2)

### Serological testing and data processing

Based on a review of the published literature on immune responses to *V. cholerae* infection [30–32], we selected seven known cholera-related antigens to investigate with a multiplex bead assay. These included O1 serogroup Ogawa serotype O-specific polysaccharide (OSP, part of the LPS), O1 serogroup Inaba serotype OSP, cholera toxin B subunit (CT-B), cholera toxin holotoxin (CT-H), Toxin co-regulated pilus subunit A (TcpA), *V. cholerae* cytolysin (VCC) (also known as hemolysin A), and *V. cholerae* sialidase. Additionally, O139 serogroup OSP (*V. cholerae* O139 serogroup is rarely detected as circulating in the last decade, but is included in the most commonly used oral cholera vaccines), heat labile enterotoxin subunit B (LT-B), and heat labile holo-enterotoxin (LT-H) (expressed during infection with enterotoxigenic *Escherichia coli* [ETEC] and have a high degree of homology with cholera toxin counterparts) and influenza haemaglutinin 1 (Flu) (as a control antigen) were also selected. Antigens were produced as previously described [33–38], or purchased from a commercial source, and are described in detail in the testing protocol. OSP antigens were produced as OSP:BSA (bovine serum albumin) conjugates to facilitate binding to the polysterene beads [33–35]. All antigens were conjugated to Luminex magnetic beads using carbodiimide coupling according to the manufacturer’s recommendations.

Each plate included a dilution series (from pooled convalescent sera of 5 patients with culture-confirmed *V. cholerae* O1 infection) and control wells, all of which were run in triplicate. Following the testing protocol, serum, beads, and secondary antibodies binding to IgG, IgA, and IgM (Southern Biotech, Catalog Numbers 9040, 2050 & 9020) were added to each well. Samples were run on a Luminex Flexmap 3D machine at Massachusetts General Hospital by one technician. Bead counts and median fluorescence intensity (MFI) values were exported from the Exponent software program. Plates were retested when over half of the positive control dilutions had >=5 antigens with a coefficient of variation (calculated from triplicate MFI measurements) greater than 20%.

For the analysis, any measurements with a bead count less than 30 were excluded (<0.1%). MFI values were averaged across replicate wells. We standardized MFI values from the assay to help adjust for inter-plate variability by calculating the relative antibody unity (RAU) (Supplementary Methods) [39]. For each plate, we fit a 4-parameter log-logistic model to the dilution series and used the median of parameter estimates to predict the RAU for each sample. (Figure S3, S4, S5, and S6) [40,41]. For samples with a predicted RAU outside the range of 10^5^ and 10^2^, the RAU was set at the threshold value. Additionally, we calculated the Net MFI for each sample (i.e., MFI of sample - MFI of blank well, but censored at 10 FI units). Despite some between-plate variability and limits of detection, we observed high correlation between the Net MFI and RAU measurements, across time points. Thus, we conducted all primary analyses using RAU with corresponding Net MFI-based results presented in the supplement.

### Statistical Analysis

We fit hierarchical regression models for each marker to estimate the degree of antibody boosting post-infection and its decay rate after the boost for each serological marker. We used a Bayesian framework with two components: a kinetic model and a measurement model (Supplementary Methods, [42]). For the kinetic model, we assumed individuals had a linear rise in log concentration of antibodies from 5 days post-infection to an individual-specific peak, followed by exponential decay over time. In the measurement model, we assumed random error was normally distributed (on the log-scale) and accounted for the fact that some observations were censored (e.g., vibriocidal titers and RAU measurements at the interpolation boundaries). We fit the models using Markov Chain Monte Carlo methods implemented in Stan [41]. We also estimated the association of age group (<10 years vs. ≥10 years), sex (male vs. female), blood type (O blood type vs. non-O blood type), and infecting serotype (Ogawa vs. Inaba), on baseline antibody levels, boosting, and decay. We also fit biphasic decay models and compared with the exponential parameterization using the loo package [43].

We explored the ability of statistical models to identify individuals who were recently infected with *V. cholerae* O1. We used four definitions of recent infection (i.e. infection windows): infections having occurred (i) 5-45 days, (ii) 5-120 days, (iii) 5-200 days and (iv) 5-300 days before blood collection. Measurements from uninfected household contacts and cases infected <5 days prior (due to insufficient time to generate an antibody response) were always considered as not recently infected. Using serological biomarkers and three non-immunological demographic variables (age, sex, and blood type), we trained random forest classification models to identify recently infected individuals [44]. We removed 11 (4%) of samples (from cases) for this analysis as they were either missing a vibriocidal or ELISA measurement. We fit models with weights to account for both class imbalance and for the large concentration of measurements collected during the early convalescent period (7-30 days) compared to later post-infection period (Figure S7, Supplementary Methods). Using 10-fold cross-validation, we estimated the cvAUC to evaluate the ability of the model to identify recently infected individuals. We kept all measurements from each individual within the same cross-validation fold. To understand which markers had the largest influence on model fits, we used a permutation importance metric [45]. To understand how our model choice may have impacted our results, we fit three alternative models (i.e. Lasso and Elastic-Net Regularized Generalized Linear Models, Bayesian Additive Regression Trees, and Extreme Gradient Boosting) and combined results from each to yield ensemble predictions and an accompanying cvAUC [46].

We also evaluated the specificity and time-varying (i.e., time since infection) sensitivity of the random forest classification models using leave-one-individual-out cross-validation. For each fold (i.e. left-out individual), we fit random forest models to 100 random samples of 50% individuals in the training set and found a cut-off that satisfies the Youden Index or a desired specificity cut-off (90%, 95%, or 99%) in the other 50% of individuals. Using the median value of these cut-offs and a model fit with the entire training data, we predicted the serostatus of the left-out samples. Using the predicted serostatus for each sample from leave-one-individual-out cross-validation, we fit hierarchical logistic regression models to estimate the specificity and time-varying sensitivity of each random forest model [47]. For time-varying sensitivity models, we assumed that the logit(sensitivity) was a function of (log-transformed) days since infection [48]. We allowed for increasingly complex functions as time since infection increased, including a constant sensitivity for the 45-day model, a linear decrease in sensitivity for the 120-day model, a quadratic polynomial for the 200-day model and a cubic polynomial for the 300-day infection window model.

Data and code used to select samples and conduct primary analyses are available at: https://github.com/HopkinsIDD/cholera-multiplex-panel. The lab protocol for the MBA assay is available at dx.doi.org/10.17504/protocols.io.3byl4b1x8vo5/v1. Original data collection was approved by the icddr,b Ethics Review Board and these analyses were deemed exempt from review by the Johns Hopkins Bloomberg School of Public Health Institutional Review Board.

## Results

### Description of individuals and timing of samples

We tested 296 samples from 48 confirmed cholera cases (4 to 1,083 days post infection) and 9 samples from 3 uninfected household contacts of cases (Table S1). *V. cholerae* serogroup O1 was isolated from each case with most being serotype Ogawa (81%) and the rest being serotype Inaba. The median age of cases at time of enrollment was 11 years (interquartile range (IQR): 6-26 years) with 17% being <5 years old and 35% being ≥18 years old. Most cases were male (62%) and nearly half had the O blood type (46%).

All cases had a baseline sample collected <5 days after infection. Nearly all cases had additional samples collected between 6-11 days (n=46), 28-36 days (n=46), 87-110 days (n=42) and 172-191 days (n=39) post symptom onset. Between 268 and 1083 days after infection, 1 person had three samples, 35 people had two samples, 2 people had one sample, and 10 people had zero samples collected (Figure S2).

### Kinetics of biomarkers in confirmed cholera cases

After infection, the levels of several *V. cholerae* O1-specific antibodies in most cases had a steep rise followed by variable decays (Figure 1, Figure S8). Robust anti-CT-B, anti-CT-H, anti-Inaba OSP, and anti-Ogawa OSP antibody responses were observed across the study cohort (except anti-CT-B and anti-CT-H IgM). Individuals who had an increase in anti-sialidase, anti-TcpA, and anti-VCC antibodies tended to be adults. Among anti-CT-B, anti-CT-H, anti-Inaba OSP and anti-Ogawa OSP antibodies, the observed median day of peak measurement was 8 days for IgA, between 8 and 63 days for IgM, and 25 to 33 days for IgG. As expected, there were no substantial increases in anti-O139 OSP or anti-Flu antibodies after infection (Figure S8, Figure S10). Some antibody responses were highly correlated due to antigen homology, including cross-specificity between the similar Ogawa and Inaba OSP antigens, and the CT and LT antigens [31] (Figure S11).

**Figure 1:**
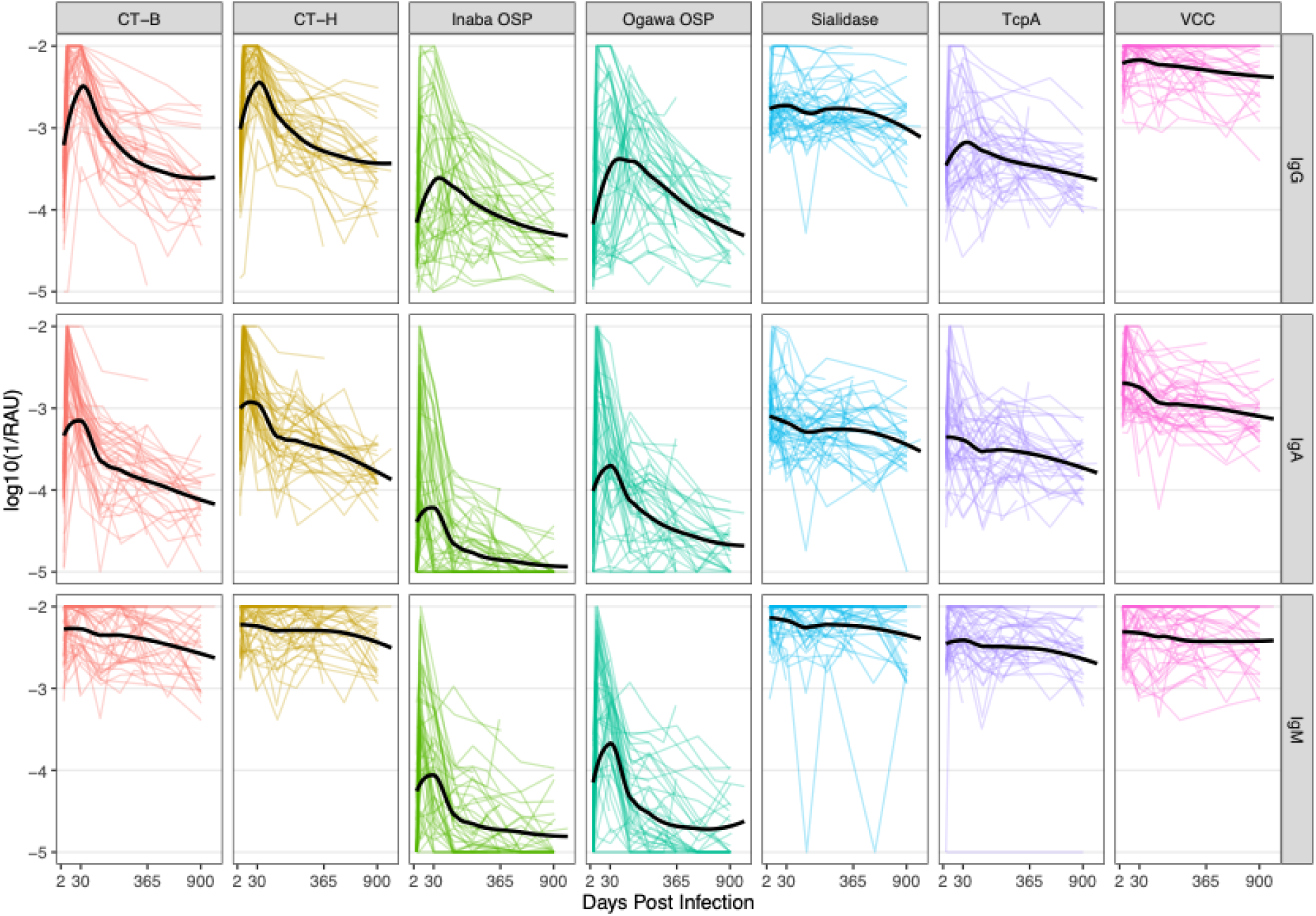
Antibody concentrations of IgG, IgA, and IgM against *V. cholerae O1* antigens among culture confirmed cholera patients. The x-axis is square-root transformed. Each colored line indicates individual trajectories over time. The Black solid line is a loess smooth function. A similar plot with Net MFI can be found in Figure S8. Trajectory plots of other measured antigens can be found in Figure S9 and Figure S10.

We fit a series of statistical models to estimate the population-level rise and decay of each marker. As biphasic models did not consistently fit better than exponential models (i.e., only in 12 of 39 markers did biphasic models fit significantly better [Table S2]), we assumed that antibody decay was exponential. These models were able to reproduce individual-level antibody trajectories (Figure S12-S17) well. The magnitude of antibody rise and duration of half-life varied considerably across MBA measures of antibody levels and vibriocidal titers (Figure 2, Table S3). The fold-increase in vibriocidal titers (45 average fold-rise for Ogawa and 44 for Inaba) was higher than for all MBA-measured antibodies except anti-Ogawa OSP IgM. We estimated relatively large increases in anti-Ogawa OSP and anti-Inaba OSP antibodies across isotypes (range: 17-69 average fold-rise) and large increases in anti-CT-B and anti-CT-H IgG and IgA antibodies (range: 22-34 average fold-rise). Anti-Ogawa OSP IgG antibodies had the longest estimated half-life (122 days [95% CI: 91-165]), similar to that of vibriocidal Ogawa antibodies (118 days [95% CI: 74-201]). Anti-CT-B IgG, anti-CT-H IgG, anti-Inaba OSP IgG, anti-TcpA IgG, and vibriocidal Inaba antibodies had slightly shorter half-lives (60-84 days) while the half-life for IgA and IgM antibodies was generally shorter (range: 3-51 days). Antibodies measured by ELISA had less pronounced responses (average boosts were all <10-fold) and were relatively short lived (half-lives were all <51 days) (Table S3).

**Figure 2:**
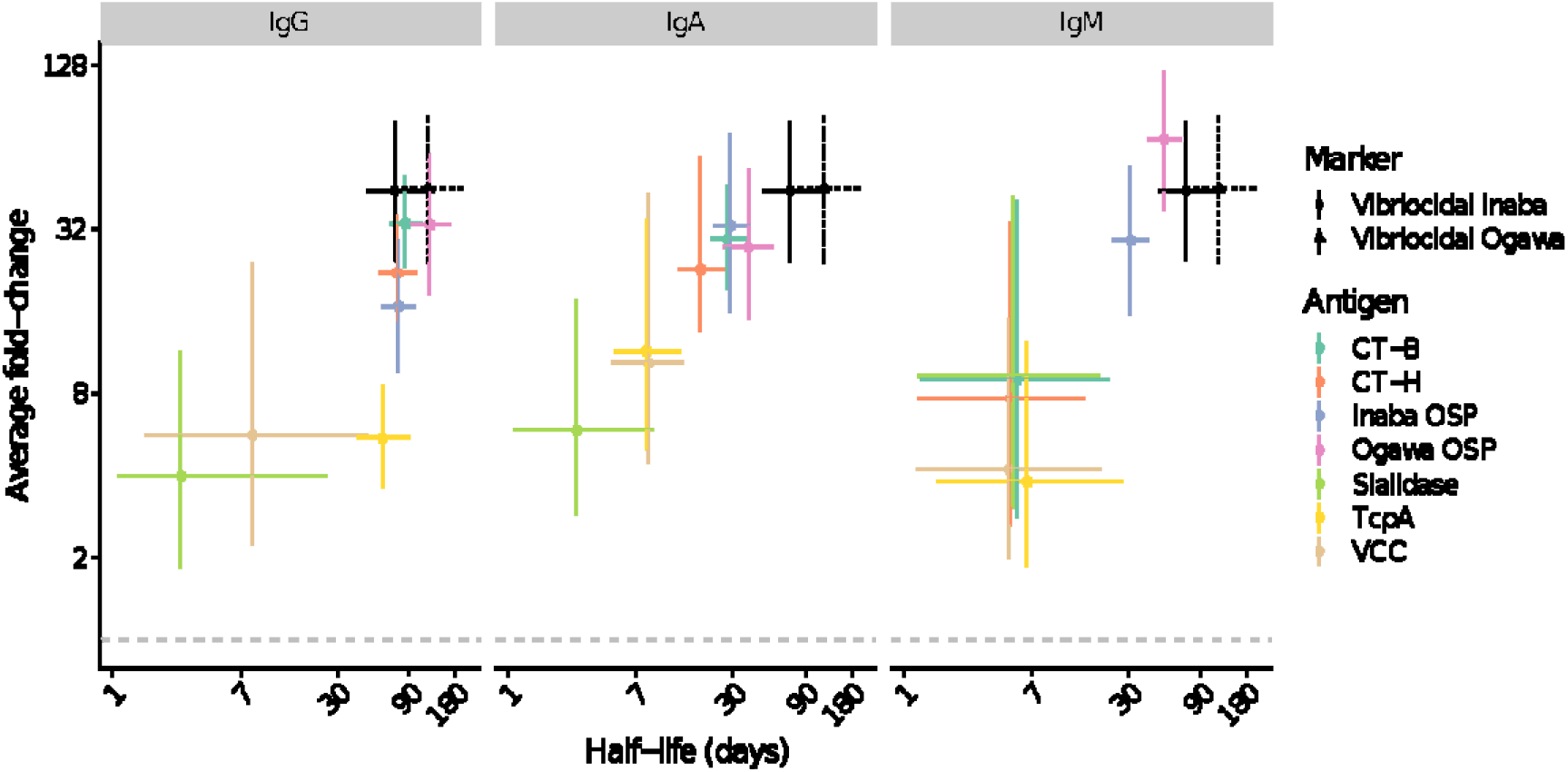
Estimated duration of half-life and average fold-change from exponential decay models. Each point indicates the median estimate of the average individual fold-rise from baseline to peak (y-value) and the median estimate of the half-life (x-value) for exponential decay univariate models. Marginal 95% credible intervals are shown as lines. Model estimates for the vibriocidal assay are shown for reference and are identical across panels. A similar plot with Net MFI can be found in Figure S20.

Both age and infecting serotype influenced antibody kinetics (Figure S18, Figure S19). We found that individuals <10-years old tended to have smaller anti-Ogawa and Inaba OSP IgG boosts (i.e. fold-increases were 3.6 (95% CI 1.2-10.6) and 3.4 (95% CI 1.2-9.3) times smaller) followed by slower decay (i.e. difference in half-life was 172 days (95% CI 70-319) and 64 days (95% CI -10-173) longer) compared to those ≥10-years old (Table S4). Individuals <10-years old had lower baseline and boosts for anti-Ogawa and anti-Inaba OSP IgA, but the rates of decay were similar. Individuals <10-years old had little difference as compared to those ≥10-years old in their anti-OSP IgM and anti-CT-B trajectories for any isotype. Individuals ≥10-years old had an increase in anti-TcpA IgG that was 2.2 times higher (95% CI 1.0-4.8) than those <10-years old. Individuals with Ogawa infections on-average had 7.2 (95% CI 2.0-24.3) times higher increases in anti-Ogawa OSP IgM, 3.6 (95% CI 1.0-12.3) times higher increases in anti-Ogawa OSP IgG, and 2.7 (95% CI 0.7-10.4) times higher increases in anti-Ogawa OSP IgA than those with Inaba infections (Table S4). On average, increases in anti-Inaba OSP IgG, IgA, or IgM were similar regardless of infection serotype. There were little differences in terms of baseline values, boost, or decay rates by O-blood group or sex.

### Identification of recent infections with cross-sectional serologic measurements

We estimated receiver operator curves and their accompanying cross-validated area under the curves (cvAUCs) from random forest models trained on 18 MBA markers (three isotypes and six antigens) and three individual non-immunological factors aimed at identifying recently infected individuals; measurements of anti-CT-H antibodies were not included in these models given their high correlation with that of anti-CT-B antibodies. The average cvAUC was consistently above 89% regardless of infection window but was higher at shorter time windows (Figure 3A, Figure S21). Models using observations that were weighted based on time since infection either performed equally or slightly worse than models without weights. As an ensemble of four different machine learning models performed only slightly better than the random forest, with a great increase in complexity, we conducted further analyses with the random forest model alone (Figure S22).

**Figure 3:**
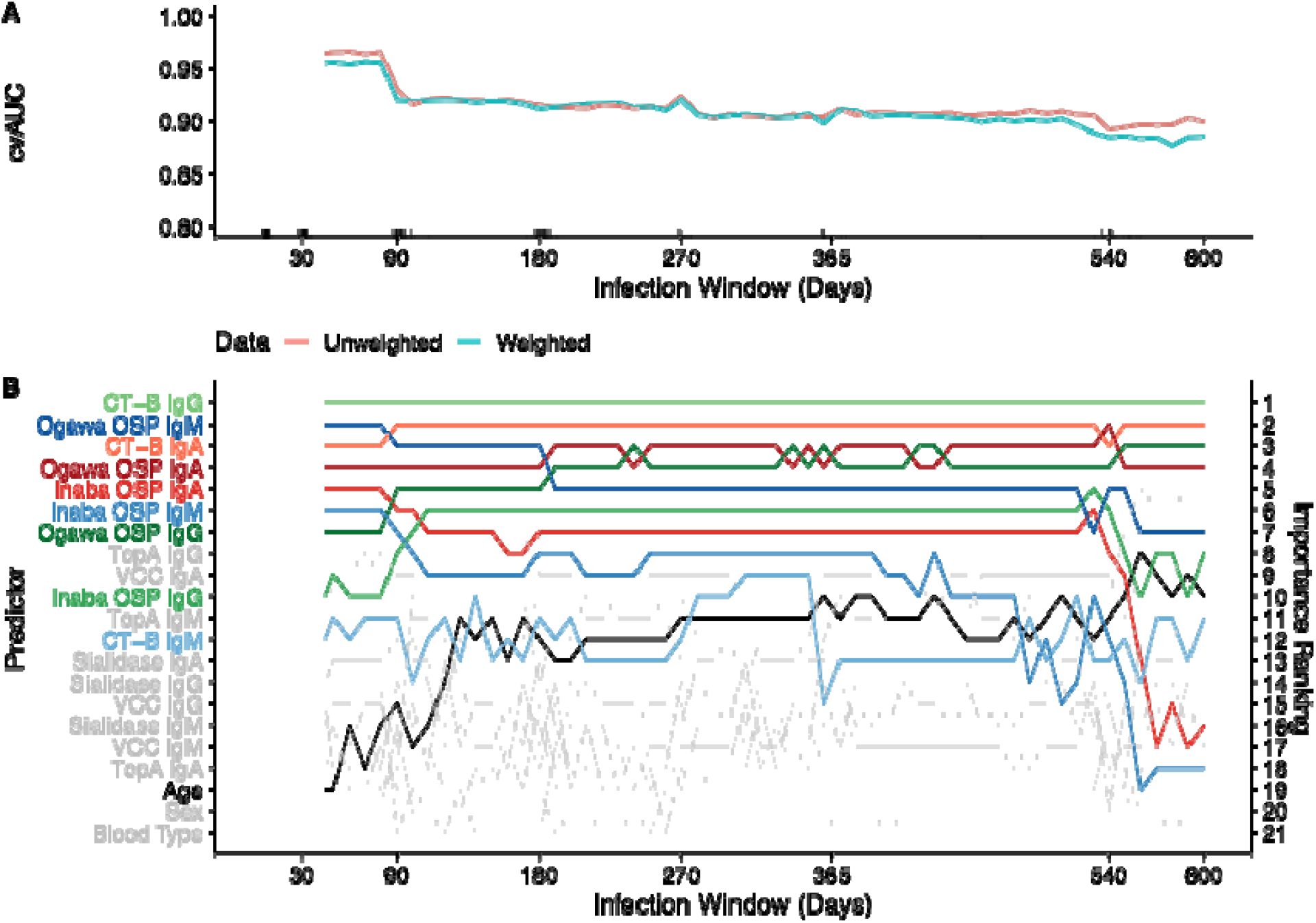
Cross-validated area under the receiver operating characteristic curve (cvAUC) and predictor importance rankings for MBA markers across random forest models with varying infection windows. Estimates of mean cvAUC (10-fold) and 95% confidence interval are shown for weighted and non-weighted models between 50- and 600-day infection windows at 10-day intervals (A). Rug plot shows the day of collection of samples from cases used in training models. Samples collected under 5 days since infection, over 600 days since infection, or from household contacts are not shown. For each infection window of weighted models, the rankings of predictors by their importance are shown on the y-axis (B). Colors of lines are unique to each predictor. A similar plot with Net MFI can be found in Figure S23.

Across infection windows, anti-CT-B IgG antibodies were consistently the most influential predictor of recent infection while the relative importance of predictors changed with different infection windows (Figure 3B, Figure S21). With infection windows shorter than 90 days, anti-Ogawa OSP IgM was the second most influential marker, but waned in influence over longer windows, while the relative influence of anti-Ogawa OSP IgG increased over time. Anti-CT-B IgA and anti-Ogawa OSP IgA were consistently among the most influential markers. Anti-Ogawa OSP markers were always more influential than anti-Inaba OSP markers within isotype (likely because 81% of cases used to train the models were infected with the *V. cholerae* Ogawa serotype). Other antibodies, age, sex, and blood type did not greatly influence the model.

We then compared cvAUC from models fit to MBA measurements with those fit with the traditional vibriocidal and ELISA measurements for four infection windows (45-day, 120-day, 200-day and 300-day). The model fit with both vibriocidal and ELISA markers was highly predictive of recent infection at 45-day (cvAUC: 97%), 120-day (cvAUC:92%), 200-day (cvAUC:88%) and 300-day (cvAUC: 88%) infection windows (Figure 4A, Table S5). The cvAUC of models trained with all 18 MBA markers was consistently similar to models trained with vibriocidal and ELISA markers (range of cvAUC ratios: 0.99-1.05). (Figure 4A, Table S5)

**Figure 4.**
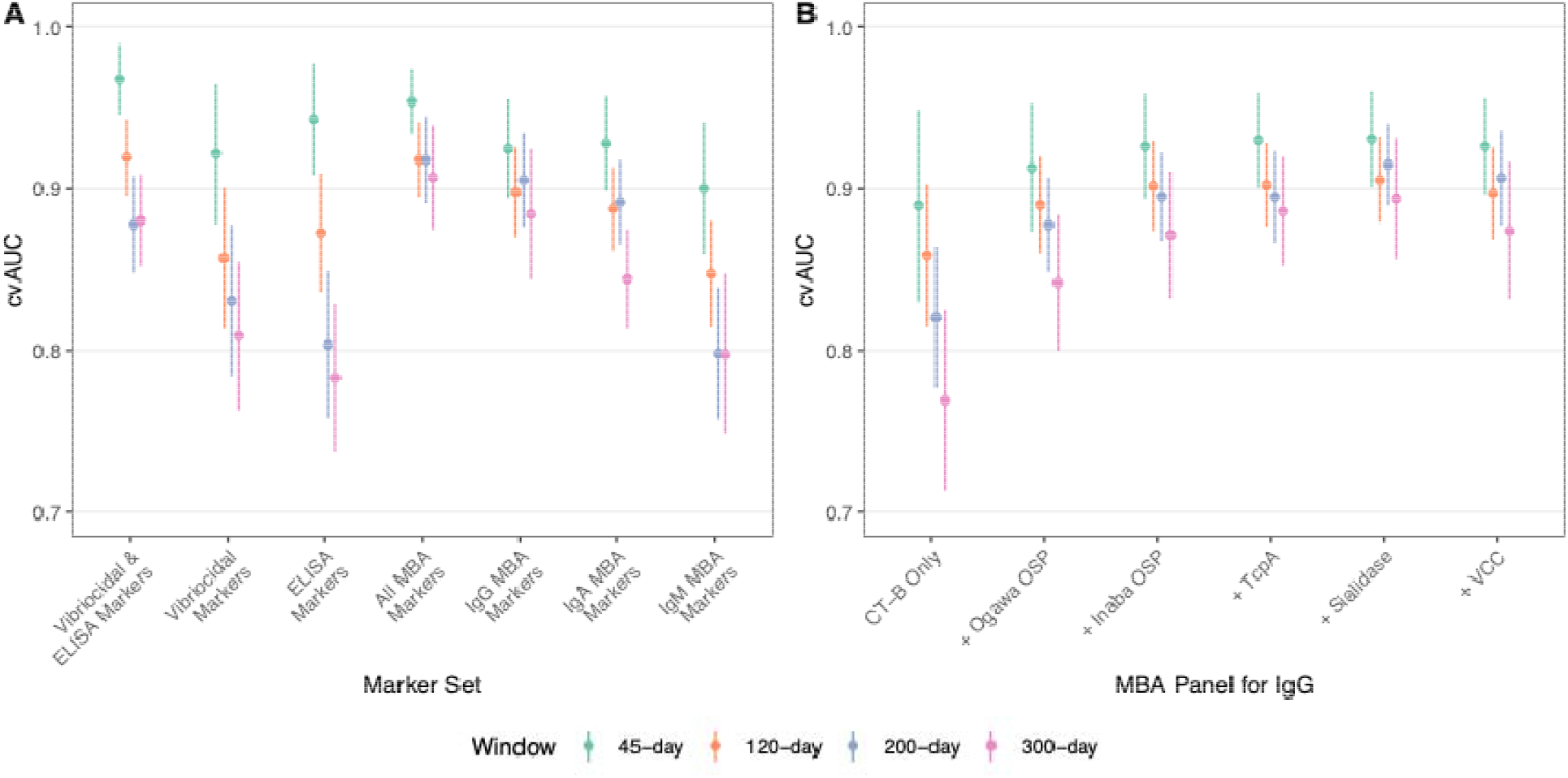
Comparison of cross-validated AUC across random forest models trained on traditional and MBA serological markers for 45-day, 120-day, 200-day, and 300-day infection windows. Random forest models were fit using a specified marker set and individual level factors including age, sex, and blood type (A). Estimated mean and 95% confidence intervals for cvAUC are reported. Models fit to reduced panels of IgG MBA markers are shown (B). The order of how antigens were added was determined by the variable importance when fitting a model with only IgG MBA markers. A similar plot with Net MFI can be found in Figure S24.

### Simplifying the multiplex bead assay panel

We explored how using fewer MBA markers would impact model performance (Figure 4, Table S5). Across all timescales, a model using six IgG MBA markers (e.g., 200-day cvAUC: 91%, 1% lower mean cvAUC, Figure 4B) was slightly more predictive than those using all 6 IgA MBA markers (e.g., 200-day cvAUC: 89%, 3% lower mean cvAUC) or all 6 IgM MBA markers (e.g., 200-day cvAUC: 80%, 12% lower mean cvAUC).

As many commonly used serosurveillance panels are based on IgG markers alone, [49] we considered a reduced panel with only IgG (Figure 4B, Table S6, Figure S22). A model using only anti-CT-B IgG and non-immunological predictors was predictive (200-day cvAUC: 82% [95% CI: 78%-86%]) of recent cholera infection across all infection windows. Adding both Ogawa OSP and Inaba OSP led to additional improvement (200-day cvAUC: 89% [95% CI: 87%-92%]). The addition of TcpA, VCC and Sialidase had little impact on the overall cvAUC (200-day model: 91% [95% CI: 88%-94%]). Model without age, sex, and blood type had similar performance to their counterparts with these non-immunological predictors.

We then estimated the specificity and time-varying sensitivity of random forest models fit with traditional and MBA markers using leave-one-out cross-validation (Figure 5). When using the Youden Index (i.e., jointly maximizing sensitivity and specificity), median estimates of specificity were all below 90% (Figure 5A). Sensitivity estimates were negatively correlated with specificity estimates and did not vary substantially between models with different marker sets when using the same infection window (Figure 5B). When fixing specificity at 90%, the (non-time-varying) median sensitivity estimates ranged from 71% to 90% across different models for the 45-day infection window. As for other infection windows, sensitivity steadily decreased over time for the 120-day (range of median estimates: 35%-98%), 200-day (range of median estimates: 21%-93%), and 300-day (range of median estimates: 8%-92%) window.

**Figure 5.**
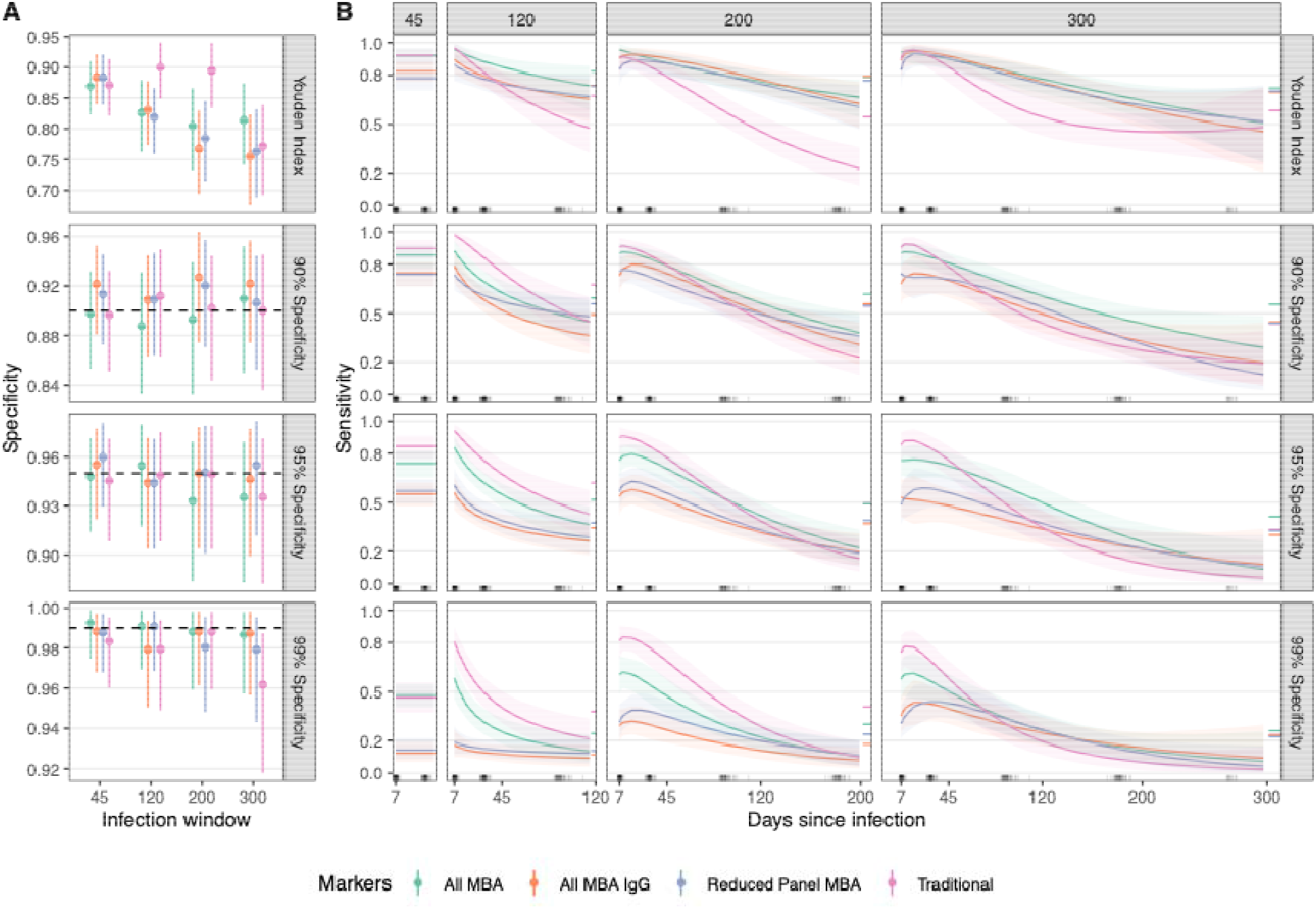
Specificity and time-varying sensitivity estimates of random forest models trained with leave-one-out cross-validation for 45-day, 120-day, 200-day, and 300-day infection windows using different cut-offs. Median and 95% credible intervals are shown for the estimated (A) nominal specificity (black dashed line) and (B) time-varying sensitivity. Each row represents a different method for acquiring a cut-off including the Youden Index or maximizing sensitivity for a desired value of specificity. The relationship between logit(sensitivity) and time since infection (log-transformed) was constant for the 45-day window, linear for the 120-day, quadratic for the 200-day window, and cubic for the 300-day window. Traditional = vibriocidal Ogawa, vibriocidal Inaba, and 4 ELISA markers, All MBA = 18 MBA markers, All MBA IgG = 6 MBA markers, Reduced panel= Ogawa OSP, Inaba OSP, and CT-B IgG. All models also included age, sex, and blood type as predictors. A similar plot with Net MFI can be found in Figure S25.

## Discussion

We developed an MBA to measure antibody responses to *V. cholerae* infection and then modeled the utility of these measures to estimate seroincidence. When measured by MBA, patients with cholera developed anti-OSP and anti-CT-B antibody increases that were of higher magnitude and duration than responses measured by ELISA and were comparable in magnitude and duration to traditional vibriocidal titers. Models using MBA-measured antibodies accurately identified individuals infected within 300 days before blood collection, and were equally accurate as models combining vibriocidal assays with ELISAs. Owing to its accuracy and scalability, the use of a *V. cholerae*-antigen MBA provides an opportunity to increase global cholera seroincidence data and can be incorporated in multi-pathogen serosurveillance systems without using a combination of traditional serologic assays.

We also measured responses to a suite of potentially informative antibody responses that have not been assessed for their utility in estimating seroincidence. These markers included VCC, sialidase, and TcpA. Compared to OSP- and CT-responses, antibody responses to VCC, sialidase, and TcpA were highly variable and on aggregate were of lower magnitude and short duration. These features limit the utility of these novel biomarkers for estimating disease incidence. However, this may have been due to the large number of children in our sample, who compared to adults, had a less robust antibody response to these antigens (Figure S11). This is consistent with previous studies showing that repeated exposures to *V. cholerae* are required to consistently produce antibodies to TcpA [36] and that in an endemic area responses to *V. cholerae* sialidase are associated with increasing age [38].

Given the lack of consistent and durable responses to VCC, sialidase, and TcpA, it is not surprising that a simplified MBA assay, measuring only responses to CT-B and OSP performed similarly to a full suite of antigens. Taken together, these results suggest that with the increasing use of multi-pathogen integrated serosurveillance using MBAs, the inclusion of three additional antigen targets (CT-B, Ogawa OSP, and Inaba OSP) to larger panels could efficiently provide data on *V. cholerae* O1 infection rates. While many serosurveillance panels only measure IgG antibodies [49], advancements to simultaneously measure multiple isotypes could further improve detection of recent infections, especially when seroincidence over a short infection window is of interest [50].

Our study has some limitations. First, we analyzed data from a cohort of patients with severe cholera in a highly endemic area. Many infections with *V. cholerae O1* lead to mild disease and these infections may lead to different post-infection antibody responses [51,52], potentially leading to misclassification. Despite the small number of individuals included in this study; our samples were selected to be ‘representative’ of a larger previously published cohort (Supplement) thus enhancing the generalizability of our findings. Individuals in Bangladesh are also likely infected several times throughout their lives [48] with each successive infection acting to boost antibody levels to higher levels than would be observed in an otherwise immunologically naïve population. Reassuringly, however, we previously demonstrated that models fit to traditional markers on Bangladeshi patients performed well when used in North American challenge study volunteers [19]. Finally, the extent to which vaccinated individuals may be classified as recently infected using this model is not known. As cholera vaccines are used more frequently in cholera endemic settings, further studies are needed to optimize seroincidence models in partially vaccinated populations. For MBA-based platforms in which additional antigens and isotypes can be readily added, the addition of biomarkers that distinguish vaccination from infection could be a critical component of estimating infection incidence and, when combined with other data, disease burden.

As large investments in cholera prevention and control measures are being made, serosurveillance is likely to be an important tool for tracking trends in incidence to better target interventions and measure their effectiveness in reducing infections. We show that measuring responses to as few as three antibodies with MBA can identify individuals infected up to one year before, with similar precision as traditional serologic methods which rely on less scalable functional measures of immunity. While cholera specific panels may be warranted in some locations, inclusion of *V. cholerae* specific beads in larger multi-pathogen MBAs being used across the world could lead to a better understanding of the epidemiology of cholera and improve our access to key data to aid the fight against cholera.

## Data Availability

Data and code used to select samples and conduct primary analyses are available at: https://github.com/HopkinsIDD/cholera-multiplex-panel. The lab protocol for the MBA assay is available at dx.doi.org/10.17504/protocols.io.3byl4b1x8vo5/v1.

## Acknowledgments

We thank Stephen Lauer for his initial analyses in the selection of serum samples for MBA testing and the Infectious Disease Dynamics Group at Johns Hopkins University for the feedback and advice on the statistical analysis. We are grateful to Dr. Slavomír Bystrický, Institute of Chemistry, Slovak Academy of Sciences, Bratislava, Slovak Republic for the provision of *V. cholerae* O139 OSP. We acknowledge the study participants and families who consented to enroll into this study.

