## Supplemental Materials for "Identifying recent cholera infections using a multiplex bead serological assay"

2022-06-27

### Supplementary Methods

#### Method S1: Overview of sample selection

##### Prediction-based sampling

We aimed to select a sample of the original cohort data from Bangladesh (the SMIC and PIC cohorts) to test out for this analysis of a multiplex bead assay. Rather than attempt to handpick individuals using descriptive guidelines, we decided to choose the sample that best predicts the rest of the cohort. The sample with the best prediction accuracy should have attributes that reflect that of the whole cohort, be they demographic (age, sex, blood type) or test-based (bactericidal, ELISAs).

The process for making these predictions was as follows:

1. Choose a random sample of individuals to create the “training set”, for a given scenario (explained below)
2. Fit a random forest model to those individuals to classify individuals as recently infected or not
3. Create a “balanced test set” (explained below)
4. Use the random forest to predict the outcomes in the balanced test set
5. Evaluate the predictions with cross-validated area under the ROC curve (cvAUC)

We looked at three scenarios for selecting the training set, all of which included 20 individuals who are under 10 (‘children’) and 20 individuals who are 10 and older (‘adults’). The scenarios only differ by the number of household contacts used; it is important to note that all household contacts in PIC/SMIC were adults. In the first scenario, we randomly draw 10 adults who were index cases and 10 adults who were household contacts. In the second scenario, we draw only index cases and no household contacts. In the third scenario, we draw from the pool of index and household contacts indiscriminately; letting the predictive ability of the models choose the number of household contacts.

After selecting the training set and fitting the random forest model, we predicted a “balanced test set”. The test set consisted of the observations that were not chosen in the training set. We took a large random sample with replacement of the test set, but balanced such that (a) there were as many seropositive (as predicted by the model) observations as seronegative observations and (b) the seropositive observations were evenly distributed over their time since infection. This is important because the original cohort was skewed towards having more seropositive observations with lower times since infection. Having an equal number of seropositive and seronegative cases ensured that we did not reward models for erring on the side of making positive or negative predictions. By sampling across time since infection, we hoped to improve the model performance at times further from infection; the serosurvey model performed quite well at short time intervals but poorly at longer horizons.

After resampling 10,000 times, we chose a set that had the highest cvAUC for the third scenario. This included 38 cases and 2 household contacts.

##### Sample inventory

After investigation of freezer stocks, samples from 39 of 40 individuals were found. Samples from two additional cases were added to the set, giving a selection of 245 samples from 41 individuals (39 cases and 2 household contacts).

##### Additional sample selection

After initial testing was completed, we decided to supplement the dataset with samples from 10 individuals (9 cases and 1 contact) in order to balance out the ages of individuals. In particular, we were concerned about the lack of individuals with samples between the ages of 10 and 18 in the original sample. To limit the influence of boosted antibody responses from reinfection/exposure during the follow-up period, we removed any data points that were part of or after a greater than a 2 fold-rise between measurements in vibriocidal Ogawa titers >90 days post initial infection. Our final sample included 305 samples from 51 individuals (48 cases and 3 contacts).

#### Method S2: Procedure for calculating the relative antibody unit

On each 384-well plate, a dilution series of positive control wells was included to adjust for between-plate variability. We used this to calculate the relative antibody unit (RAU) for all samples on the plate. The RAU is the expected dilution of the positive control sample needed to get the same MFI as the sample.

##### Dilution series

Serum samples from 5 patients with culture-confirmed *V. cholerae* O1 collected 7 days after symptoms were combined to create a serum pool. Of the total 19 plates run, 7 had four dilution points (1:100, 1:400, 1:1,600, 1:6,400). After deciding to expand the range of dilutions, the next 12 plates had eight dilution points (1:10, 1:40, 1:160, 1:640, 1:2,560, 1:10,240, 1:40,960, 1:163,840). Additionally, blank control wells (i.e. not including any sera) were run on every plate. All samples were run in triplicate and MFI values were averaged.

##### Model parameterization and fit

For each antigen, we fit four-parameter log-logistic models to each plate’s dilution series. As previously described (1), the relationship between dilution and MFI is defined as follows:

$$\begin{matrix} Y_{i} & =\text{log(median fluorescence intensity) for sample }i \\ x_{i} & =\text{dilution for sample }i \\ Y_{i} & =c+\frac{d-c}{1+exp\left( b\times\left( log\left( x_{i} \right)-log\left( e \right) \right) \right)} \end{matrix}$$

We chose this parameterization given its frequent use for modeling dose-response relationships and that we only had four dilution points for many of our plates. For plates with seven dilution points, we used the drc package in R (1). For plates with four dilution points, we decided to implement a Bayesian framework so that we could fit the model with prior distributions informed by the seven dilution series wells:

$$\begin{matrix} b & \sim\text{Normal}\left( \mu_{b},1 \right) \\ c & \sim\text{Normal}\left( \mu_{c},0.5 \right) \\ d & \sim\text{Normal}\left( \mu_{d},20 \right) \\ e & \sim\text{Normal}\left( \mu_{e},100 \right) \end{matrix}$$

Specifically, $\mu_{c}$, the mean of the prior distribution for $c$ (the lower bound of the logistic curve), was set equal to the log(MFI) for the blank sample for each plate. The variance parameter of the prior distribution for $c$ was set at 0.5 to be slightly larger than observed among blank samples. The mean values for the other priors ($\mu_{b}$,$\mu_{d}$, and$\mu_{e}$) were set equal to the average values estimated from the seven dilution models. Variance values for the prior distributions of $b$,$d$,and $e$ were selected to be sufficiently large to be relatively uninformative.

##### Relative antibody unit calculation

For each antigen on each plate, the mean value for each parameter was used to calculate the relative antibody unit (RAU). To avoid extrapolation, all samples with a calculated RAU above 1:100 or below 1:100,000 were set equal to those values. Any samples with MFI values falling outside of the logistic curve were also set to the threshold value.

#### Method S3: Univariate decay models

We fit univariate decay models to describe the antibody dynamics of individuals for each biomarker. All models had two components: 1) a measurement model and 2) a decay model. This data set contains $n$ individuals (indexed by $i$), and $m$ total measurements (indexed by $j$). Each marker is modeled separately.

##### Measurement Models

We had several different types of measurements of biomarkers (multiplex bead assay (MBDA) RAU, Vibriocidal titers, and ELISA concentrations) to fit models to. We assumed that the observed data were normally distributed ($f_{N}$ denotes the normal density) around the true (unobserved) value. Each different model accounted for censored observations in various ways based on how the data was generated.

- $Y_{ij}$ = the true value at the time of the $j$th measurement for individual i.
- $Y_{ij}^{*}$ = the observed value at the time of the $j$th measurement for individual i.
- $\sigma$ = measurement error variance

###### Multiplex bead assay relative antibody unit measurement model

For our analysis, the RAU was inverted and log10 transformed. Values can only take a value above -5 and below -2. Any value set at -5 and -2 are censored values and are treated as so:

$$Pr\left( Y_{ij}^{*}|Y_{ij},\sigma\right)=\left\{ \begin{matrix} \int_{-\infty}^{-5} f_{N}\left( Y_{ij}|\sigma\right) & \text{if }Y_{ij}^{*}=-5 \\ f_{N}\left( Y_{ij}|\sigma\right) & \text{if }-5<Y_{ij}^{*}<-2 \\ \int_{-2}^{\infty} f_{N}\left( Y_{ij}|\sigma\right) & \text{if }Y_{ij}^{*}=-2 \end{matrix} \right.$$

###### Vibriocidal assay measurement model

Vibriocidal titers were divided by 5 and log2 transformed. The vibriocidal assay outputs a measurement of the highest dilution where the vibriocidal reaction still occurs. Therefore, the true dilution is between the reported dilution and the next dilution (i.e. interval censored). $V_{max}$, the largest measureable log titer, was 11. $V_{min}$, the smallest measureable log titer was 0.

$$Pr\left( Y_{ij}^{*}|Y_{ij},\sigma\right)=\left\{ \begin{matrix} \int_{-\infty}^{V_{min}} f_{N}\left( Y_{ij}|\sigma\right) & \text{if }Y_{ij}^{*}=V_{min} \\ \int_{Y_{ij}}^{Y_{ij}+1} f_{N}\left( Y_{ij}|\sigma\right) & \text{if }V_{min}<Y_{ij}^{*}<V_{max} \\ \int_{V_{max}}^{\infty} f_{N}\left( Y_{ij}|\sigma\right) & \text{if }Y_{ij}^{*}=V_{max} \end{matrix} \right.$$

###### ELISA measurement model

ELISA measurements were all log10 transformed. No values were at the limit of detection so censoring was not accounted for.

$$\begin{matrix} Pr\left( Y_{ij}^{*}|Y_{ij},\sigma\right)=f_{N}\left( Y_{ij},\sigma\right) \end{matrix}$$

##### Kinetic Models

We also explored different modes of decay (expoenential vs. biphasic). Additionally, we also investigated differences in decay between individuals with different of demographic characteristics.

###### Shared variables and parameters

The following variables and parameters are shared across all decay models:

- $T_{ij}$ = time of sample collection post-infection for individual i and measurement j.
- $\omega_{i}$ = baseline value for individual $i$
- $\lambda_{i}$ = boost for individual $i$ (restricted to values greater than 0)
- $\tau_{i}$ = duration of linear incrase for individual $i$ (restricted to values greater than 0)
- $D$ = time between infection and start of linear increase
- $\mu^{\omega}$ = average individual’s baseline rate
- $\mu^{\lambda}$ = average individual’s boost from baseline at day D (restricted to values greater than 0)
- $\mu^{\tau}$ = average individuals log time to peak from day D
- $\Sigma$ = covariance matrix for average baseline and boost

All models follow the same general temporal pattern:

1. Individuals start at their baseline values $\omega_{i}$
2. After $D$ days, an individuals value immediately increases by $\lambda_{i}$
3. Over time, an individuals value decays

We allow for baseline values and boosts to vary between individuals, but assume the decay rate is constant.

###### Exponential decay model

For the exponential model, we assume that decay follows an exponential pattern with the decay parameter $\delta$ (restricted to be greater than zero).

$$\begin{matrix} Y_{ij} & =\left\{ \begin{matrix} \omega_{i} & T_{ij}<D \\ \omega_{i}+\lambda_{i}\times\frac{T_{ij}-D}{\tau_{i}-D} & D\leq T_{ij}<\tau_{i} \\ \omega_{i}+\lambda_{i}\times e^{-\delta\left( T_{ij}-\tau_{i} \right)} & T_{ij}\geq\tau_{i} \end{matrix} \right. \\ \left( \begin{matrix} \omega_{i} \\ \lambda_{i} \\ log\left( \tau_{i} \right) \end{matrix} \right) & \sim\text{MVN}\left( \left( \begin{matrix} \mu^{\omega} \\ \mu^{\lambda} \\ \mu^{\tau} \end{matrix} \right),\Sigma\right) \end{matrix}$$

###### Biphasic decay model

For the biphasic decay model, we assume that each marker’s value is determined by two independent components that each decay exponentially. The proportion of these two components is unknown.

- $\theta_{1}$ = decay rate for first component (restricted to be greater than zero)
- $\theta_{2}$ = decay rate for second component (restricted to be greater than zero)
- $\alpha$ = proportion of boost due to first component (restricted between 0 and 1)

$$\begin{matrix} Y_{ij} & =\left\{ \begin{matrix} \omega_{i} & T_{ij}<D \\ \omega_{i}+\lambda_{i}\times\frac{T_{ij}-D}{\tau_{i}-D} & D\leq T_{ij}<\tau_{i} \\ \omega_{i}+\lambda_{i}\left( \alpha e^{-\theta_{1}\left( T_{ij}-D \right)}+\left( 1-\alpha\right)e^{-\theta_{2}\left( T_{ij}-D \right)} \right) & T_{ij}\geq\tau_{i} \end{matrix} \right. \\ \left( \begin{matrix} \omega_{i} \\ \lambda_{i} \\ log\left( \tau_{i} \right) \end{matrix} \right) & \sim\text{MVN}\left( \left( \begin{matrix} \mu^{\omega} \\ \mu^{\lambda} \\ \mu^{\tau} \end{matrix} \right),\Sigma\right) \end{matrix}$$

###### Exponential model including individual covariates (e.g. age group)

To understand how kinetics varied by individual-level attributes, we included age (<10 vs 10+ years), sex (male vs female), blood group (O group vs. non-O group), and infecting serotype (Ogawa vs. Inaba) as binary variables in the model. These covariates were allowed to modify the initial baseline, boost and decay rate.

- $X_{i}$ = covariate of individual $i$ (e.g. 0 if < 10 years and 1 if 10+years)
- $\beta^{\omega}$ = fixed effect of covariate on baseline
- $\beta^{\lambda}$ = fixed effect of covariate on boost
- $\beta^{\tau}$ = fixed effect of covariate on time to boost
- $\beta^{\delta}$ = fixed effect of covariate on decay

$$\begin{matrix} Y_{ij} & =\left\{ \begin{matrix} \omega_{i} & T_{ij}<D \\ \omega_{i}+\lambda_{i}\times\frac{T_{ij}-D}{\tau_{i}-D} & D\leq T_{ij}<\tau_{i} \\ \omega_{i}+\lambda_{i}\times e^{-\left( \delta+\beta^{\delta}X_{i} \right)\left( T_{ij}-\tau_{i} \right)} & T_{ij}\geq\tau_{i} \end{matrix} \right. \\ \left( \begin{matrix} \omega_{i} \\ \lambda_{i} \\ log\left( \tau_{i} \right) \end{matrix} \right) & \sim\text{MVN}\left( \left( \begin{matrix} \mu^{\omega}+\beta^{\omega}X_{i} \\ \mu^{\lambda}+\beta^{\lambda}X_{i} \\ \mu^{\tau}+\beta^{\tau}X_{i} \end{matrix} \right),\Sigma\right) \end{matrix}$$

###### Calculation of key parameters

- Average fold-change: $\left\{ \begin{matrix} {10}^{\mu^{\lambda}} & \text{if MBA or ELISA} \\ 2^{\mu^{\lambda}} & \text{if Vibriocidal} \end{matrix} \right.$
- Halflife: $ln\left( 2 \right)/\delta$

#### Method S4: Sample weighting

The samples used to train random forest classification models are heavily skewed towards times in the early acute and convalescent period due to the higher density of blood draws during that period and the limited loss to follow-up early on (**Figure S1**). This skewed distribution of infection times likely does not represent the likely flatter distribution of infection times in a study population during a cross-sectional survey. We would expect that the assessments of model fit might be misleading or overly optimistic if this skew is not accounted for.

We attempted to re-weight the samples used to train random forest models in each class based on an expected distribution of time since infection. Additionally, we used weighting to account for class imbalance such that the sum of weights among recently infected and non-recently infected were made equal.

##### Expected distribution of infection times

For this analysis, we decided to assume a constant hazard of infection with an annual incidence rate of 10%. When conducting a cross-sectional serosurvey at one point in time, we would expect that the time since last infection should be exponentially distributed. This could be modified given a different understanding of distribution of last infection times. Using the cumulative distribution function of an exponential distribution, we can understand the expected proportion of samples from each time slice.

##### Defining time slices

As shown in **Figure S1**, the day of sample collection we have for each sample are clumped around certain time points. In order to properly reweight the samples we have, we need to consider what unobserved infections the samples are ‘standing in’ for.

Given a particular infection window (e.g. 200-day), we assigned each sample to whether they were inside or outside of the window. We assumed baseline samples (collected <5 days post infection) of cases and household contacts were always outside of the infection window.

Next, we further divided the time inside and outside the infection window into time slices. For cases, samples were collected around 2, 7, 30, 90, 180, 270, 360, 540, 720, 900 and 1080 days. We used these times to define the time slices using the average of consecutive time points as dividers. We assumed that samples from any case after 540 days as well as uninfected contacts belonged to the same time slice.

| 200-day Infection Window | Status | Day | Time Slice | n (%) |
| --- | --- | --- | --- | --- |
| Inside | Case | 7 | [5,18.5) | 46 (27) |
|  |  | 30 | [18.5,60) | 46 (27) |
|  |  | 90 | [60,135) | 42 (24) |
|  |  | 180 | [135,200) | 39 (23) |
| Outside | Case | 2 | [0,5) | 48 (36) |
|  |  | 270 | [200,315) | 11 (8) |
|  |  | 360 | [315,450) | 12 (9) |
|  |  | 540 | [450,Inf) | 25 (19) |
|  |  | 720 | [540,Inf) | 1 (1) |
|  |  | 900 | [540,Inf) | 25 (19) |
|  |  | 1080 | [540,Inf) | 1 (1) |
| Outside | Contact | 2 | [540,Inf) | 3 (2) |
|  |  | 7 | [540,Inf) | 3 (2) |
|  |  | 30 | [540,Inf) | 3 (2) |

All samples belonging to the same time slice are equally weighted and stand in for all potential infection times within the slice. If the proportion of samples for given time slice does not match what is expected from an exponential distribution, they can now be properly weighted.

##### Weight calculation

We calculated weights to account for both class imbalance and distribution of infection times. To account for class imbalance, we simply created weights so the sum of weights among samples inside and that of samples outside the window were equal. For the class weight, samples within each class were equally weighted.

To account for the distribution of infection times, we calculated both the observed and the expected proportion of samples to be in each time slice. The calculated time-based weight to account for infection time is the ratio of the expected versus observed proportion (**Figure S7**).

The final weight used in our analyses is the product of the class weight and the time-based weight. It was recalculated for every different length of infection window and each fold in cross-validation.

### Supplementary Figures and Tables

#### Figure S1: Vibriocidal Ogawa measurements among individuals selected from the SMIC and PIC cohorts

Vibriocidal titer measurements were transformed by dividing the measurement by 5 and then taking the logarithm (base 2) (A). X-axis indicates the approximate collection day for each measurement for cases. All measurements for uninfected household contacts are shown regardless of collection day (B).

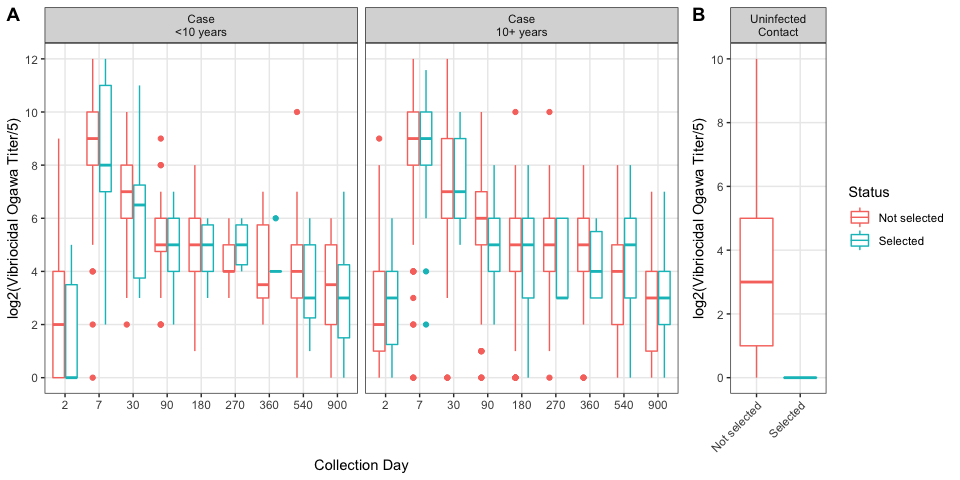

#### Figure S2: Timing of sample collection among culture confirmed cases relative to date of infection

Each point represents a blood sample collection occurred for a confirmed case (A). The daily number of samples is shown on the y-axis. Red dashed lines are placed at the threshold values for infections windows (45, 120, 200, and 300 days) (B).

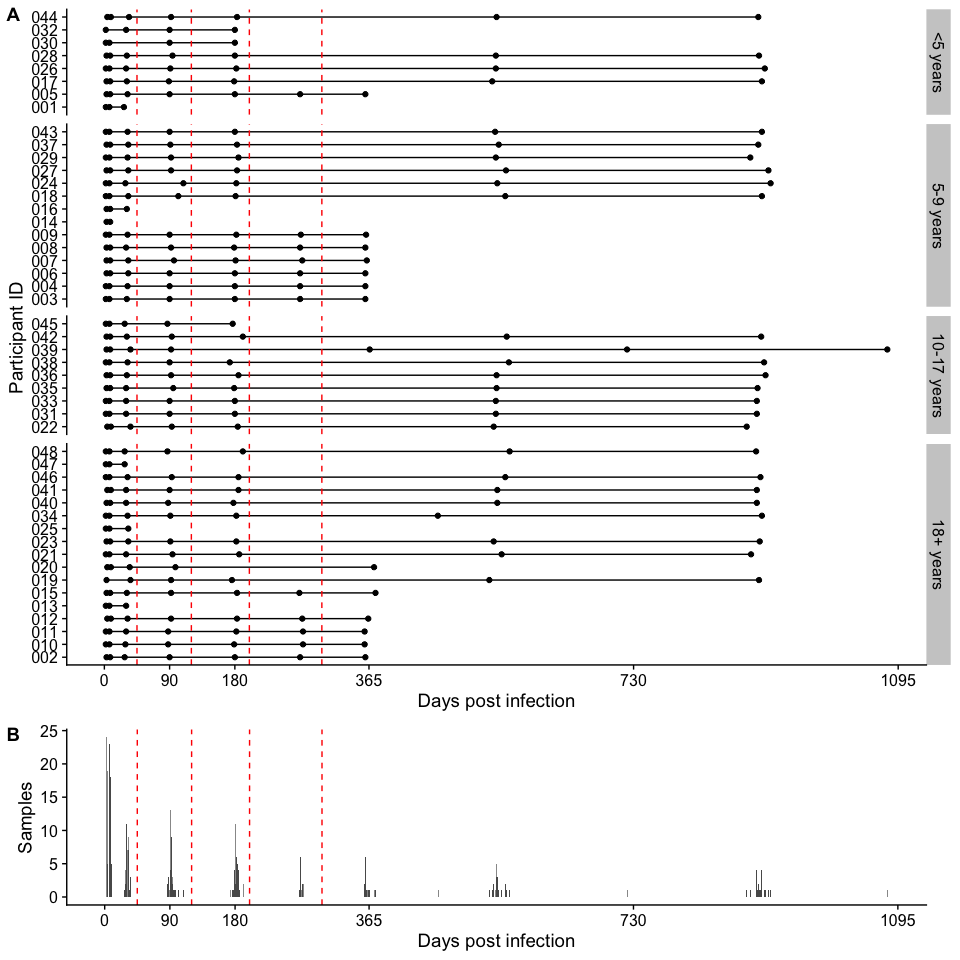

#### Figure S3: Postive control dilution curve and four parameter log-logistic curve fits for Vibrio cholerae O1 antigen multiplex bead assay markers

Each point represents the median fluorescence intensity measurement (y-value) for a dilution (x-value) of pooled convalescent sera from confirmed positive *Vibrio cholerae* O1 patients. Each plate’s dilution series were fit using a four-parameter log-logisitic regression (black lines). Shape of each point is unique to each plate.

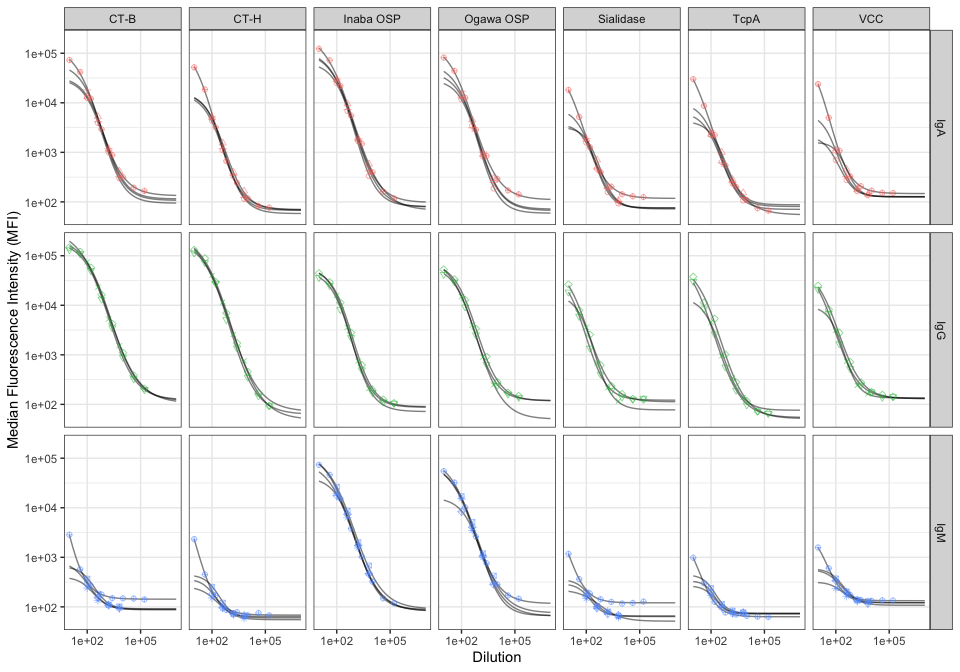

#### Figure S4: Postive control dilution curve and four parameter log-logistic curve fits for additional antigen multiplex bead assay markers

Each point represents the median fluorescence intensity measurement (y-value) for a dilution (x-value) of pooled convalescent sera from confirmed positive *Vibrio cholerae* O1 patients. Each plate’s dilution series were fit using a four-parameter log-logisitic regression (black lines). Shape of each point is unique to each plate.

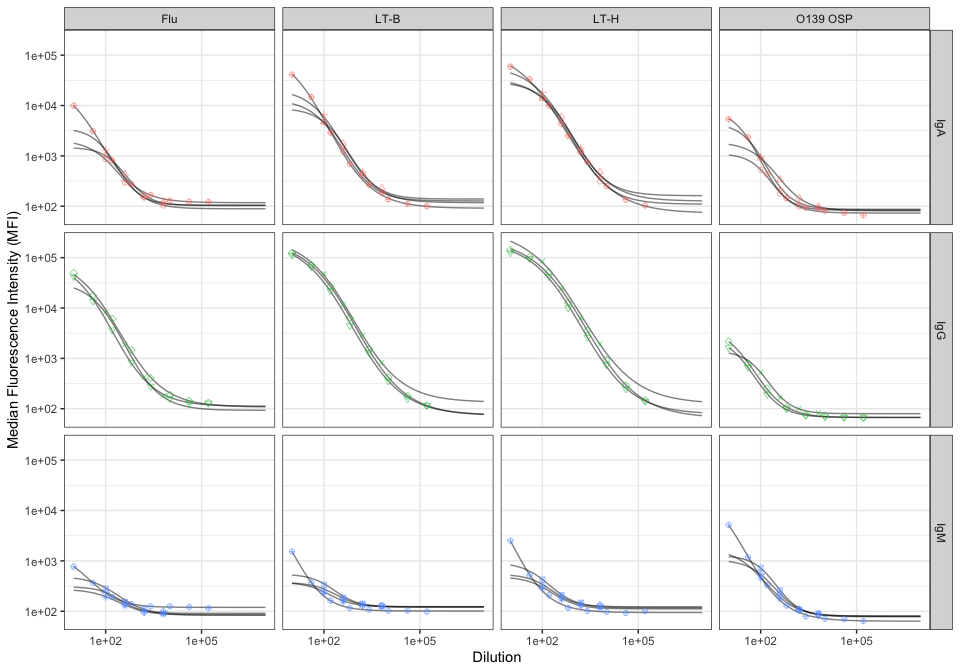

#### Figure S5: Relationship between relative antibody units and median fluorescence intensity for additional antigen multiplex bead assay markers

Relative antibody unit (RAU) measurements for each sample are plotted against the median fluorescence intensity (MFI) calculated from averaging triplicate measurements. Relative antibody units estimates were truncated at 10^2 and 10^5 (red points).

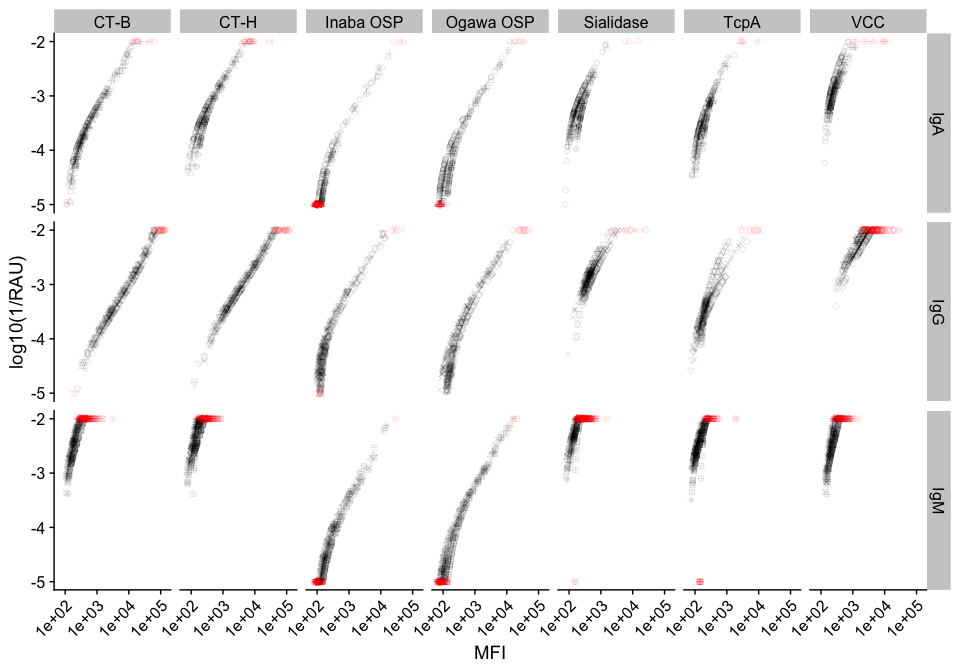

#### Figure S6: Relationship between relative antibody units and median fluorescence intensity for non-Vibrio cholerae O1 antigen multiplex bead assay markers

Relative antibody unit (RAU) measurements for each sample are plotted against the median fluorescence intensity (MFI) calculated from averaging triplicate measurements. Relative antibody units estimates were truncated at 10^2 and 10^5 (red points). LT-B = heat labile toxin B subunit. LT-H = heat labile toxin holotoxin.

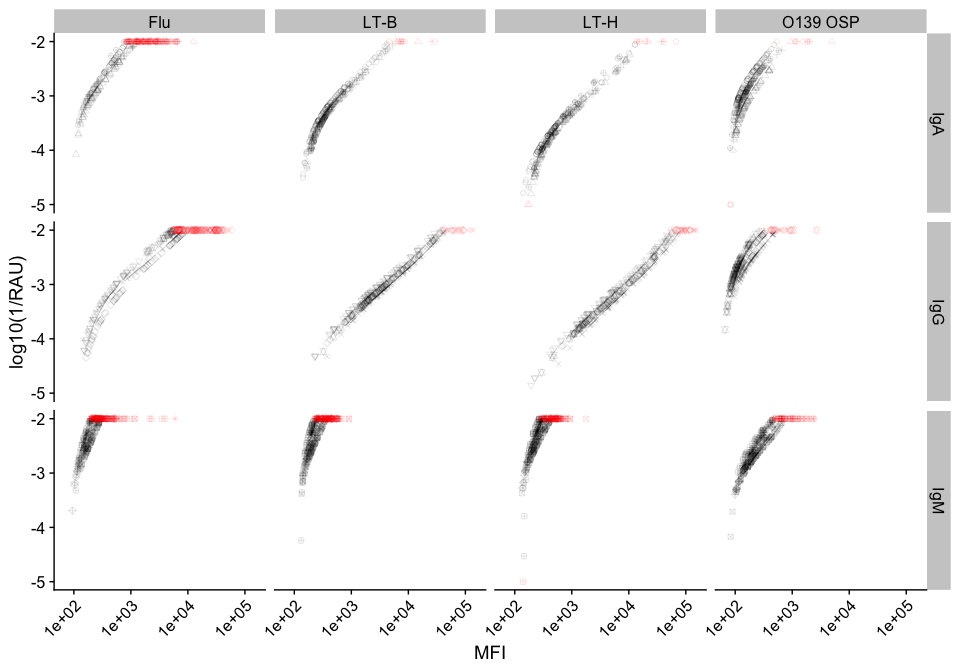

#### Figure S7: Weights used for random forest models at each infection window

The yellow columns represent the proportion of samples that fall within the time interval on the x-axis (A). The purple columns indicate the expected proportion of samples that would fall within the time interval if infection times were exponentially distributed with a 10% annual incidence. Both of these proportions condition on the sample already being either inside or outside the infection window. Each point shows the weight used for each infection window (B).

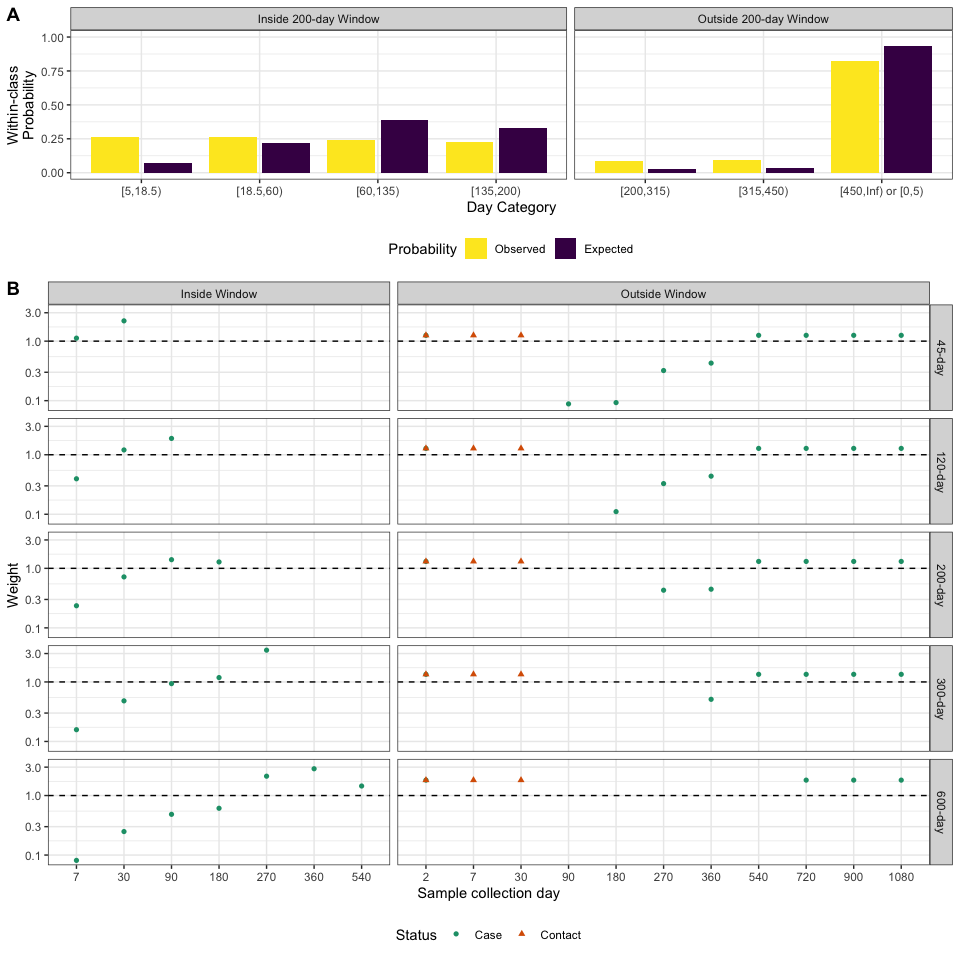

#### Table S1: Individual characteristics of culture confirmed cholera patients and uninfected household contacts

All cases were hospitalized and solely had O1 Vibrio Cholerae isolated. Inaba O1 was isolated from non-Ogawa cases

| Characteristics | Cholera cases (n=48) | Household contacts (n=3) |
| --- | --- | --- |
| Age Group |  |  |
| < 5 years (%) | 8 (17) | 0 (0) |
| 5-9 years (%) | 14 (29) | 0 (0) |
| 10-17 years (%) | 9 (19) | 2 (67) |
| 18+ years (%) | 17 (35) | 1 (33) |
| Female (%) | 18 (38) | 1 (33) |
| V. cholerae O1 Ogawa isolated (%) | 39 (81) | - |

#### Figure S8: Multiplex bead assay Net MFI measurements of IgG, IgM, and IgA against V. cholerae O1 antigens among culture confirmed cholera patients

The y-axis indicates the log (base 10) of the Net MFI and the x-axis is the number of days post-infection (square-root transformed). Each colored line indicates individual trajectories over time. The Black solid line is a loess smooth function.

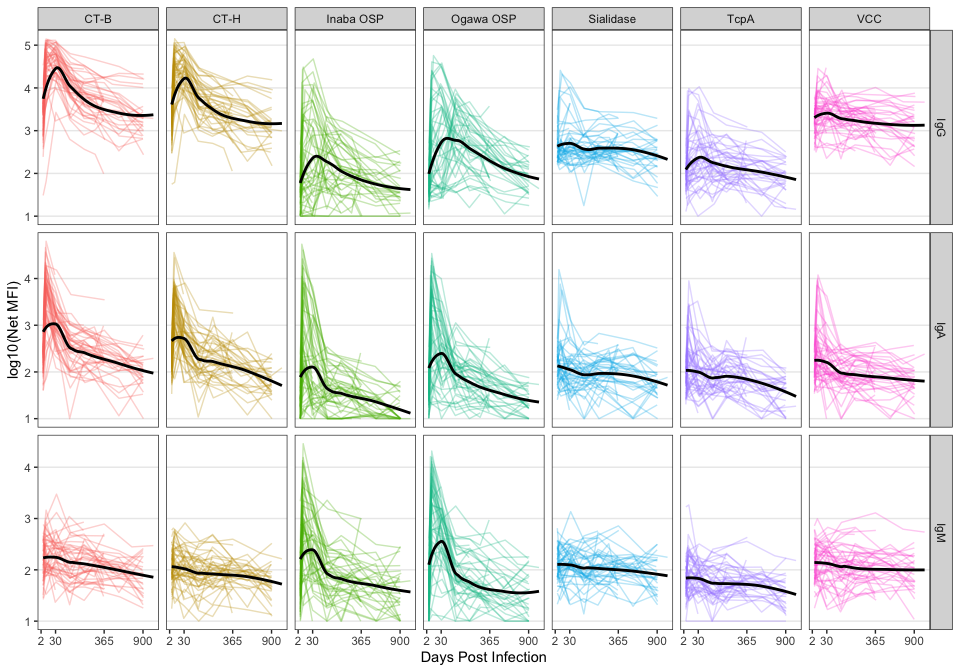

#### Figure S9: Multiplex bead assay relative antibody unit measurements of IgG, IgM, and IgA against additional antigens among culture confirmed cholera patients

The y-axis indicates the log (base 10) of the inverse RAU and the x-axis is the number of days post-infection (square-root transformed). Each colored line indicates individual trajectories over time. The Black solid line is a loess smooth function.

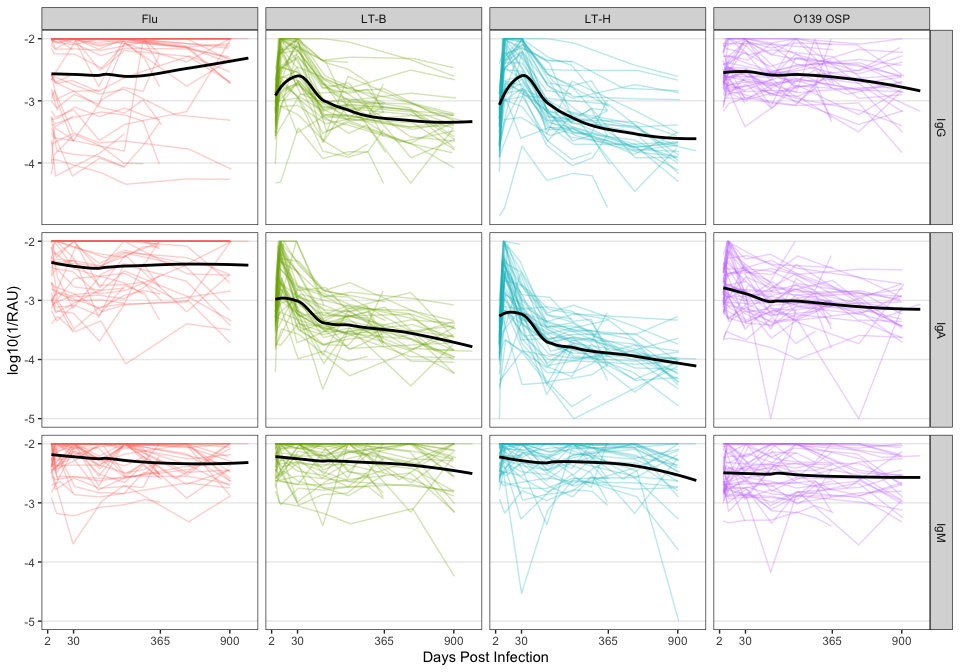

#### Figure S10: Multiplex bead assay Net MFI measurements of IgG, IgM, and IgA against additional antigens among culture confirmed cholera patients

The y-axis indicates log (base 10) of the Net MFI and the x-axis is the number of days post-infection (square-root transformed). Each colored line indicates individual trajectories over time. The Black solid line is a loess smooth function.

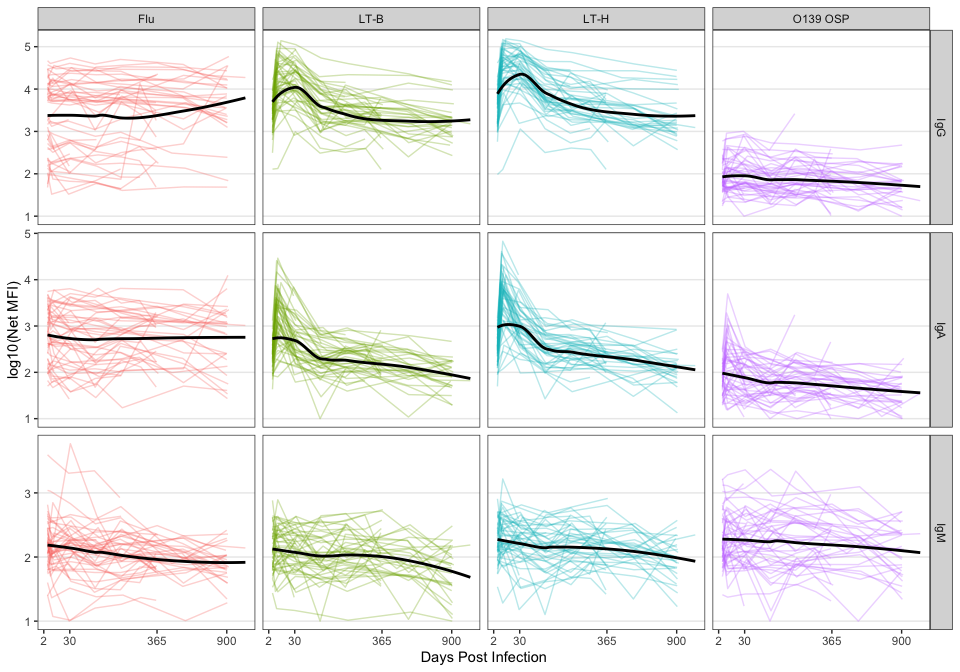

#### Figure S11: Correlation matrix of multiplex bead assay biomarkers

Spearman correlation coefficients were calculated using 49 samples all of which were collected approximately 30 days post-enrollment for both cases and contacts.

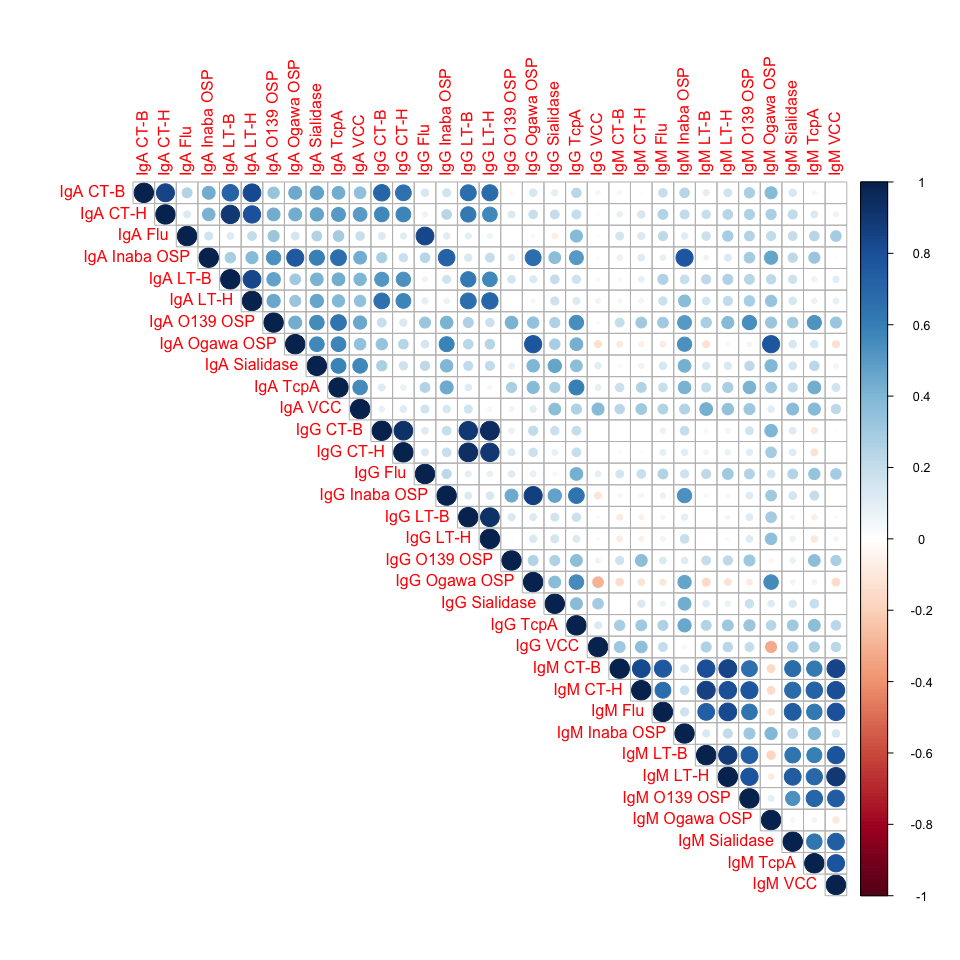

#### Table S2: Biphasic and Exponential decay model comparison

The expected log-predictive density (ELPD) and standard error difference was calculated using the loo R package. Asterisks denote that the ELPD of the biphasic model (which includes more parameters) was over 1.96 standard errors larger than the ELPD of the exponential model.See Supplementary Appendix 2 for equations defining each model

| Marker | Biphasic ELPD | Exponential ELPD | Biphasic - Exponential Standard Error Difference |
| --- | --- | --- | --- |
| ELISA CtxB IgA | -88.7 | -88.1 | -0.76 |
| ELISA CtxB IgG | 15.1 | 13.0 | 1.97* |
| ELISA LPS IgA | -132.8 | -132.5 | -1.60 |
| ELISA LPS IgG | -8.2 | -5.2 | -0.90 |
| NetMFI IgA CTHT | -126.6 | -133.8 | 3.64* |
| NetMFI IgA CtxB | -103.3 | -115.2 | 4.98* |
| NetMFI IgA Flu | -87.9 | -89.1 | 1.08 |
| NetMFI IgA InabaOSPBSA | -120.3 | -130.0 | 3.46* |
| NetMFI IgA LTB | -75.3 | -76.9 | 1.81 |
| NetMFI IgA LTh | -66.4 | -68.3 | 2.21* |
| NetMFI IgA O139BSA | -70.7 | -71.8 | 1.44 |
| NetMFI IgA OgawaOSPBSA | -149.7 | -162.6 | 4.76* |
| NetMFI IgA Sialidase | -71.0 | -71.1 | 1.77 |
| NetMFI IgA TcpA | -101.8 | -103.4 | 2.47* |
| NetMFI IgA VCC | -62.2 | -60.0 | -0.87 |
| NetMFI IgG CTHT | -102.1 | -103.9 | 2.03* |
| NetMFI IgG CtxB | -77.4 | -81.3 | 2.98* |
| NetMFI IgG Flu | -92.8 | -92.9 | 0.85 |
| NetMFI IgG InabaOSPBSA | -247.3 | -239.8 | -4.16 |
| NetMFI IgG LTB | -69.8 | -69.9 | 0.86 |
| NetMFI IgG LTh | -53.7 | -54.3 | 0.96 |
| NetMFI IgG O139BSA | -37.1 | -39.6 | 1.27 |
| NetMFI IgG OgawaOSPBSA | -177.8 | -186.7 | 5.33* |
| NetMFI IgG Sialidase | -33.8 | -32.1 | -1.29 |
| NetMFI IgG TcpA | -75.0 | -74.9 | -1.95 |
| NetMFI IgG VCC | -27.0 | -26.7 | -1.84 |
| NetMFI IgM CTHT | -6.2 | -7.8 | 0.80 |
| NetMFI IgM CtxB | -33.6 | -35.9 | 1.47 |
| NetMFI IgM Flu | -10.6 | -12.1 | 1.14 |
| NetMFI IgM InabaOSPBSA | -103.4 | -106.4 | 2.16* |
| NetMFI IgM LTB | -30.8 | -31.3 | 0.68 |
| NetMFI IgM LTh | -14.5 | -15.0 | 0.77 |
| NetMFI IgM O139BSA | -34.6 | -35.1 | 0.87 |
| NetMFI IgM OgawaOSPBSA | -113.5 | -129.0 | 5.65* |
| NetMFI IgM Sialidase | -21.5 | -21.3 | -0.56 |
| NetMFI IgM TcpA | -26.2 | -20.4 | -2.43 |
| NetMFI IgM VCC | 14.9 | 15.5 | -0.35 |
| RAU IgA CTHT | -982.0 | -536.5 | -128.71 |
| RAU IgA CtxB | -310.6 | -204.0 | -45.55 |
| RAU IgA Flu | -8,617.3 | -9,127.3 | 101.39* |
| RAU IgA InabaOSPBSA | -1,416.3 | -1,349.6 | -37.60 |
| RAU IgA LTB | -684.6 | -612.8 | -28.46 |
| RAU IgA LTh | -218.4 | -176.1 | -17.91 |
| RAU IgA O139BSA | -375.3 | -406.9 | 15.88* |
| RAU IgA OgawaOSPBSA | -246.3 | -177.6 | -33.69 |
| RAU IgA Sialidase | -121.2 | -115.3 | -3.72 |
| RAU IgA TcpA | -376.1 | -277.1 | -59.60 |
| RAU IgA VCC | -1,367.6 | -1,255.5 | -57.20 |
| RAU IgG CTHT | -979.8 | -335.8 | -167.97 |
| RAU IgG CtxB | -626.3 | -217.1 | -110.94 |
| RAU IgG Flu | -8,018.2 | -8,233.8 | 79.50* |
| RAU IgG InabaOSPBSA | -160.9 | -145.6 | -14.49 |
| RAU IgG LTB | -1,100.8 | -423.8 | -207.12 |
| RAU IgG LTh | -1,347.8 | -578.8 | -208.33 |
| RAU IgG O139BSA | -433.1 | -341.9 | -37.62 |
| RAU IgG OgawaOSPBSA | -279.4 | -199.8 | -42.23 |
| RAU IgG Sialidase | -324.8 | -347.8 | 16.85* |
| RAU IgG TcpA | -162.7 | -121.2 | -22.41 |
| RAU IgG VCC | -4,354.4 | -4,855.9 | 152.84* |
| RAU IgM CTHT | -2,319.4 | -2,383.3 | 45.56* |
| RAU IgM CtxB | -2,975.0 | -3,103.8 | 53.40* |
| RAU IgM Flu | -3,087.9 | -3,521.9 | 75.20* |
| RAU IgM InabaOSPBSA | -967.6 | -1,110.8 | 40.82* |
| RAU IgM LTB | -2,333.6 | -2,545.7 | 42.28* |
| RAU IgM LTh | -990.8 | -1,106.4 | 28.45* |
| RAU IgM O139BSA | -404.1 | -364.8 | -23.92 |
| RAU IgM OgawaOSPBSA | -546.9 | -462.0 | -23.56 |
| RAU IgM Sialidase | -1,668.8 | -1,596.8 | -46.26 |
| RAU IgM TcpA | -929.1 | -856.0 | -30.76 |
| RAU IgM VCC | -2,925.7 | -2,882.1 | -59.78 |
| Vibriocidal Inaba | -609.7 | -579.8 | -16.53 |
| Vibriocidal Ogawa | -646.3 | -595.5 | -29.28 |

#### Figure S12: Individual-level trajectories of Ogawa OSP IgG

Each facet shows the log10(RAU) measurements for individuals over time (points). Solid line indicates the median value of exponential decay model. Shaded area is the 95% credible interval.

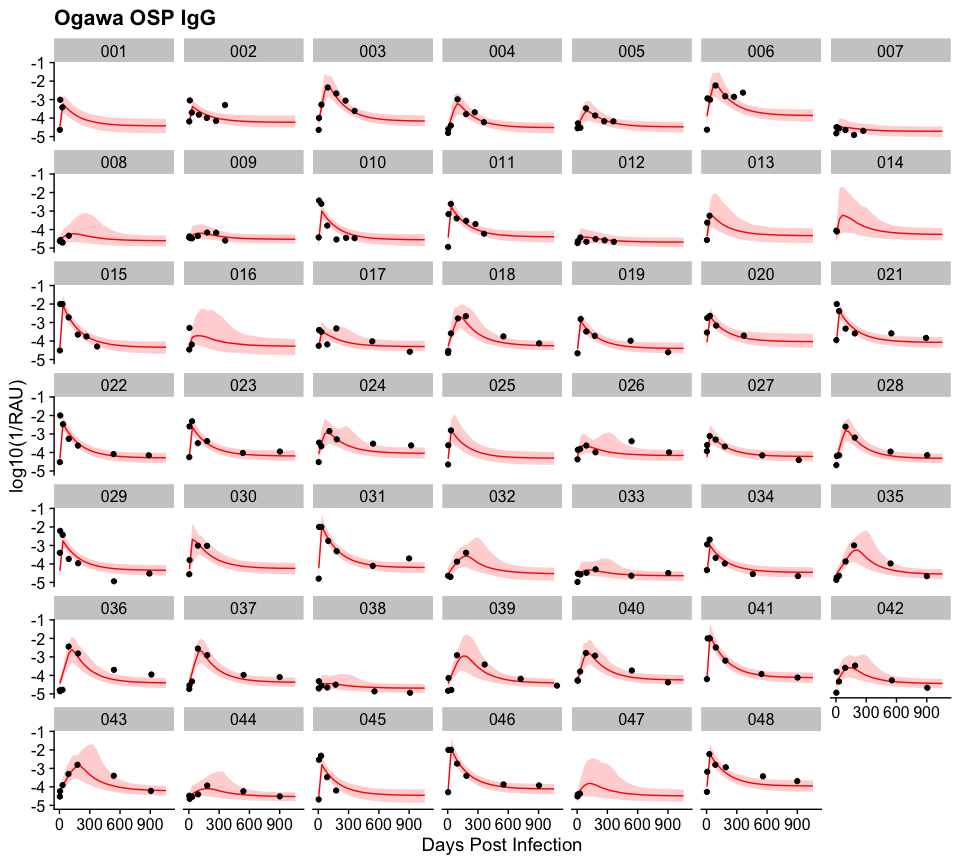

#### Figure S13: Individual-level trajectories of Ogawa OSP IgA

Each facet shows the log10(RAU) measurements for individuals over time (points). Solid line indicates the median value of exponential decay model. Shaded area is the 95% credible interval.

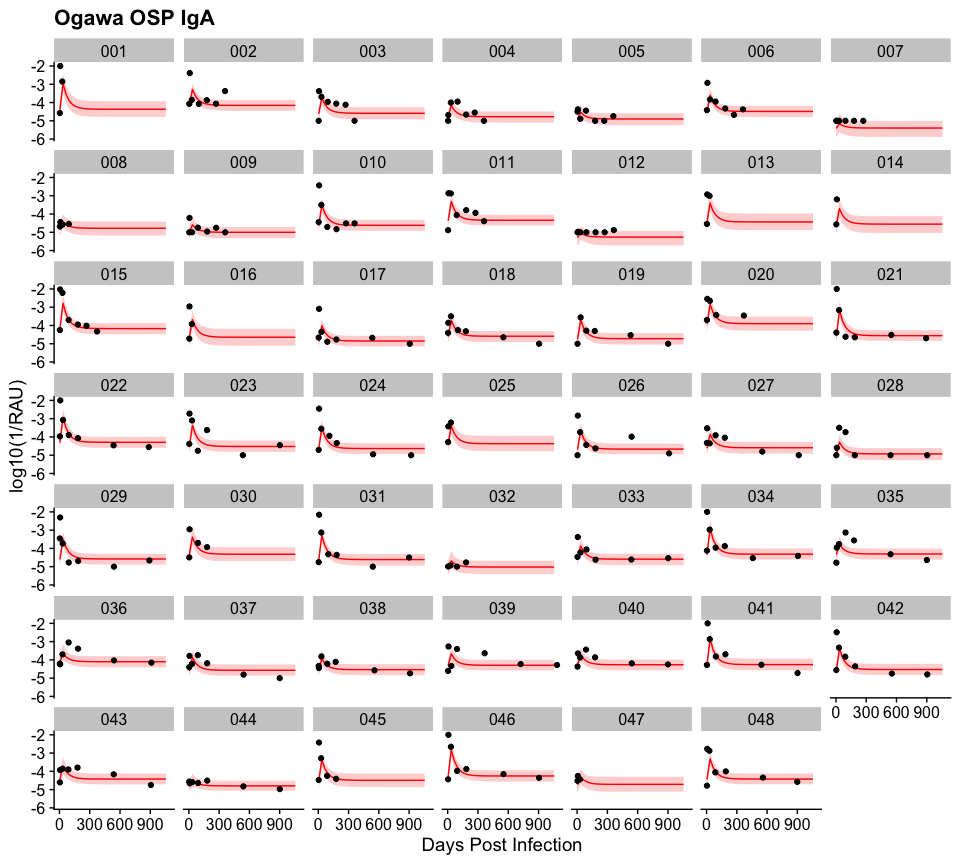

#### Figure S14: Individual-level trajectories of Ogawa OSP IgM

Each facet shows the log10(RAU) measurements for individuals over time (points). Solid line indicates the median value of exponential decay model. Shaded area is the 95% credible interval.

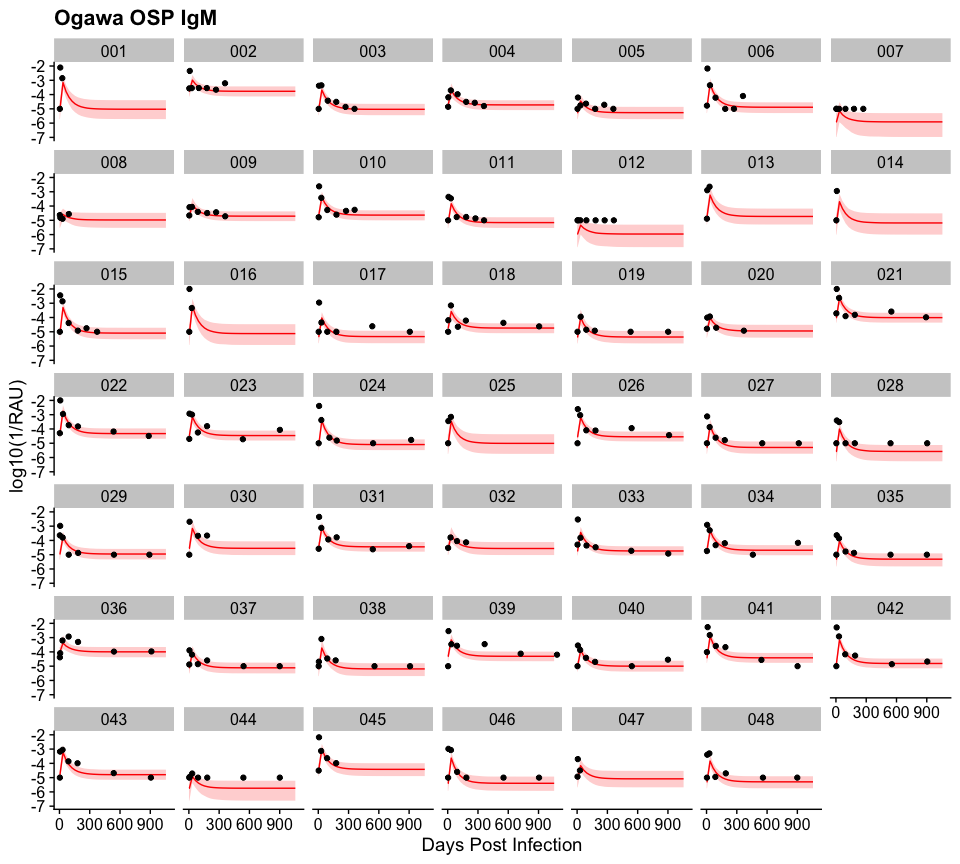

#### Figure S15: Individual-level trajectories of CT-B IgG

Each facet shows the log10(RAU) measurements for individuals over time (points). Solid line indicates the median value of exponential decay model. Shaded area is the 95% credible interval.

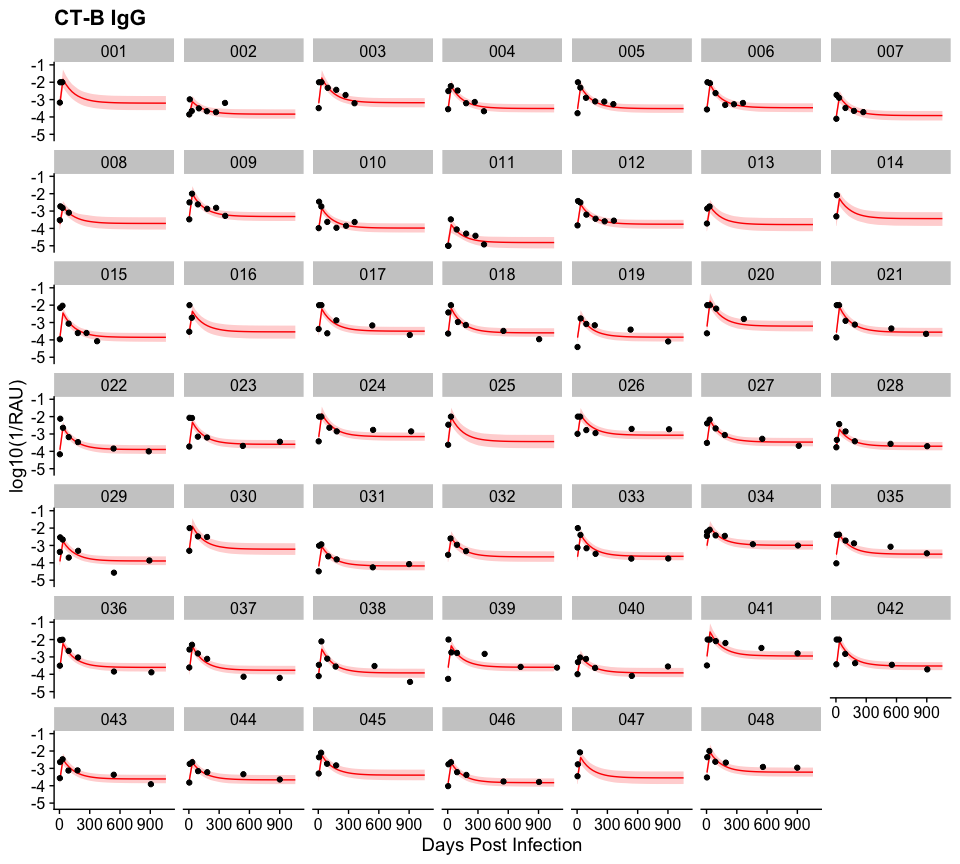

#### Figure S16: Individual-level trajectories of CT-B IgA

Each facet shows the log10(RAU) measurements for individuals over time (points). Solid line indicates the median value of exponential decay model. Shaded area is the 95% credible interval.

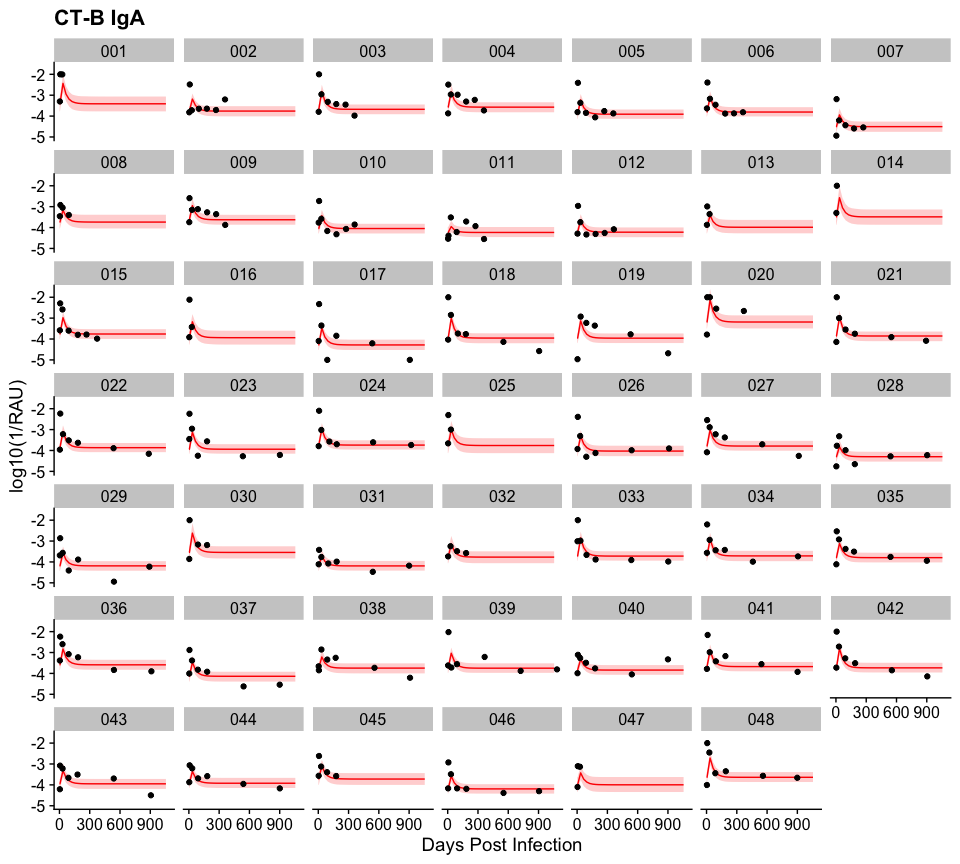

#### Figure S17: Individual-level trajectories of CT-B IgM

Each facet shows the log10(RAU) measurements for individuals over time (points). Solid line indicates the median value of exponential decay model. Shaded area is the 95% credible interval.

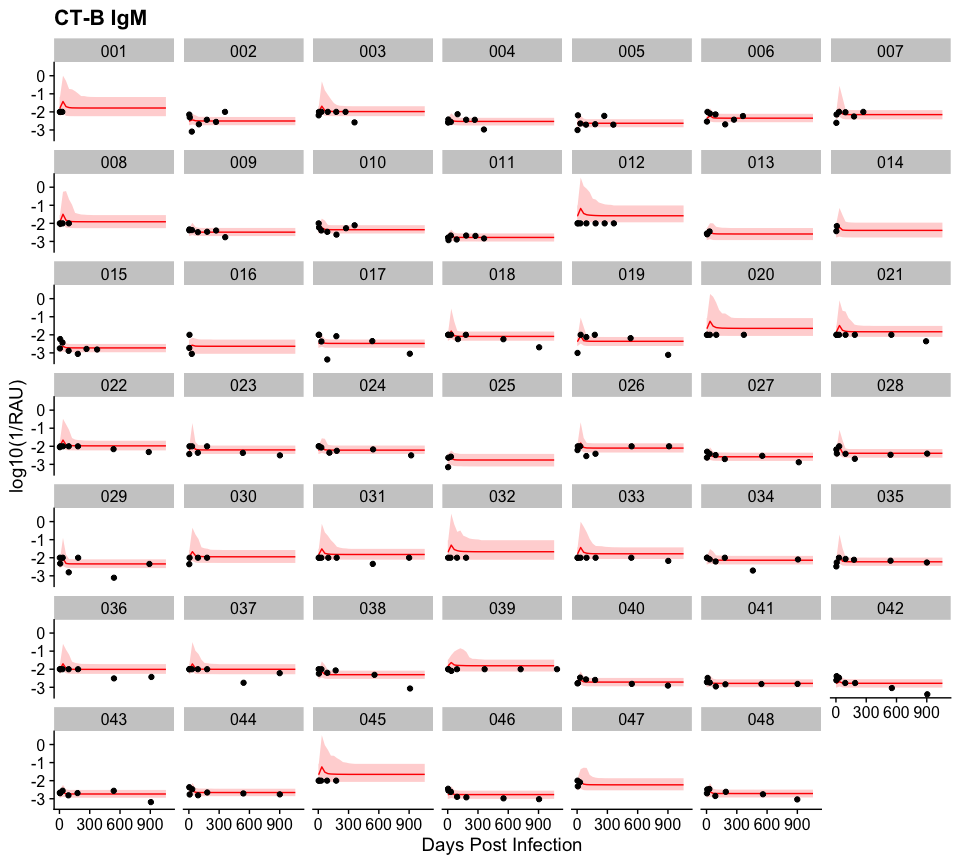

#### Table S3: Estimated duration of half-life and average fold-change from univariate exponential decay models

The median estimate duration of half-life (in days) and the median estimate increase in fold-change for the average individual are shown below as well as in Figure 2. 95% Bayesian Credible Intervals are shown in parentheses.

| Isotype | Antigen | | Half Life (95% CI) in days | | Average Fold-Change (95% CI) |
| --- | --- | --- | --- | --- | --- |
| IgA | RAU CTHT | | 18 (26-13) | | 22.9 (13.7-26.0) |
|  | RAU CtxB | | 27 (36-21) | | 29.3 (19.3-36.3) |
|  | RAU Flu | | 3 (12-1) | | 5.7 (2.0-11.6) |
|  | RAU InabaOSPBSA | | 29 (39-22) | | 33.1 (15.9-38.6) |
|  | RAU LTB | | 16 (22-11) | | 17.4 (9.7-22.5) |
|  | RAU LTh | | 21 (26-16) | | 22.6 (15.0-26.4) |
|  | RAU O139BSA | | 5 (9-1) | | 9.7 (4.1-9.3) |
|  | RAU OgawaOSPBSA | | 38 (54-26) | | 27.6 (15.2-54.2) |
|  | RAU Sialidase | | 3 (9-1) | | 5.9 (2.9-8.9) |
|  | RAU TcpA | | 8 (13-5) | | 11.4 (5.0-13.2) |
|  | RAU VCC | | 8 (14-5) | | 10.3 (4.5-13.9) |
| IgG | RAU CTHT | | 74 (98-56) | | 22.2 (14.5-97.6) |
|  | RAU CtxB | | 84 (106-67) | | 33.6 (23.5-105.8) |
|  | RAU Flu | | 3 (26-1) | | 3.1 (1.5-26.1) |
|  | RAU InabaOSPBSA | | 75 (96-59) | | 16.7 (9.6-96.0) |
|  | RAU LTB | | 46 (62-33) | | 13.1 (8.5-62.0) |
|  | RAU LTh | | 57 (71-45) | | 23.8 (15.0-70.7) |
|  | RAU O139BSA | | 4 (34-1) | | 4.1 (1.8-34.4) |
|  | RAU OgawaOSPBSA | | 122 (165-91) | | 33.2 (18.5-165.3) |
|  | RAU Sialidase | | 3 (26-1) | | 4.0 (1.9-25.6) |
|  | RAU TcpA | | 60 (89-40) | | 5.5 (3.7-88.9) |
|  | RAU VCC | | 8 (47-2) | | 5.6 (2.3-47.0) |
| IgM | RAU CTHT | | 5 (15-1) | | 7.7 (2.7-15.0) |
|  | RAU CtxB | | 5 (22-1) | | 9.0 (2.9-22.0) |
|  | RAU Flu | | 4 (22-1) | | 7.5 (2.7-22.1) |
|  | RAU InabaOSPBSA | | 31 (40-24) | | 29.1 (15.8-40.5) |
|  | RAU LTB | | 4 (15-1) | | 5.5 (2.1-14.5) |
|  | RAU LTh | | 3 (9-1) | | 6.7 (2.5-9.3) |
|  | RAU O139BSA | | 3 (149-1) | | 2.3 (1.4-148.9) |
|  | RAU OgawaOSPBSA | | 51 (66-40) | | 68.5 (38.1-66.0) |
|  | RAU Sialidase | | 5 (19-1) | | 9.3 (3.0-19.2) |
|  | RAU TcpA | | 6 (27-2) | | 3.8 (1.9-27.3) |
|  | RAU VCC | | 5 (19-1) | | 4.2 (2.0-19.5) |
| Marker | | Half Life (95% CI) in days | | Average Fold-Change (95% CI) | |
| ELISA CtxB IgA | | 16 (21-12) | | 9.5 (6.6-21.3) | |
| ELISA CtxB IgG | | 51 (75-34) | | 4.4 (3.4-75.3) | |
| ELISA LPS IgA | | 30 (46-20) | | 9.5 (6.1-45.7) | |
| ELISA LPS IgG | | 47 (74-28) | | 3.0 (2.4-74.4) | |
| Vibriocidal Inaba | | 72 (108-47) | | 44.2 (24.7-107.9) | |
| Vibriocidal Ogawa | | 118 (201-74) | | 45.2 (24.1-201.2) | |

#### Figure S18: Multiplex bead assay RAU measurements of IgG, IgM, and IgA against V. cholerae O1 antigens among culture confirmed cholera patients, by age group

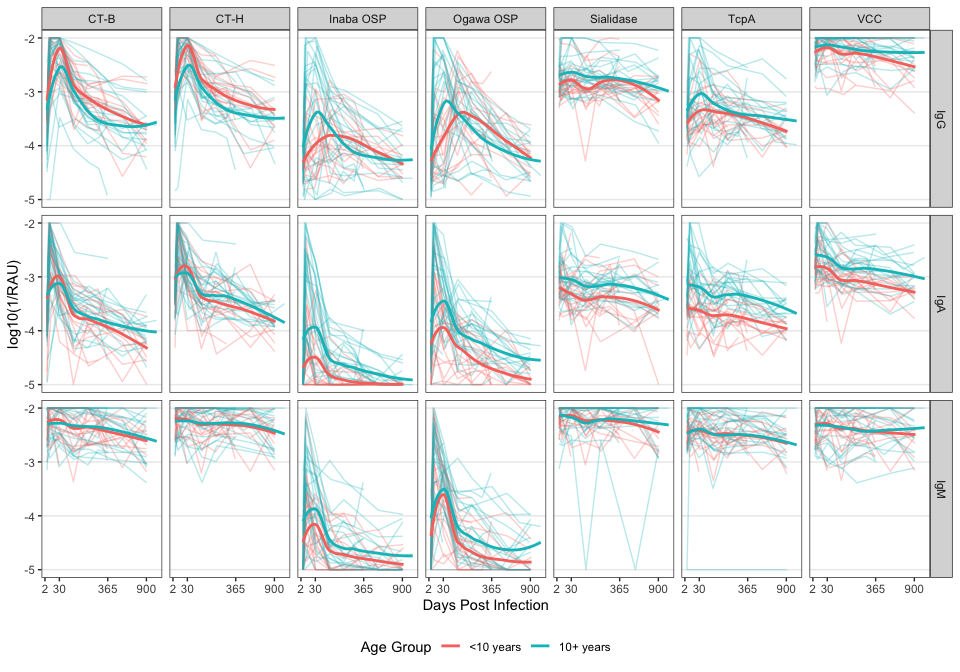

#### Figure S19: Multiplex bead assay RAU measurements of IgG, IgM, and IgA against V. cholerae O1 antigens among culture confirmed cholera patients, by infecting serotype

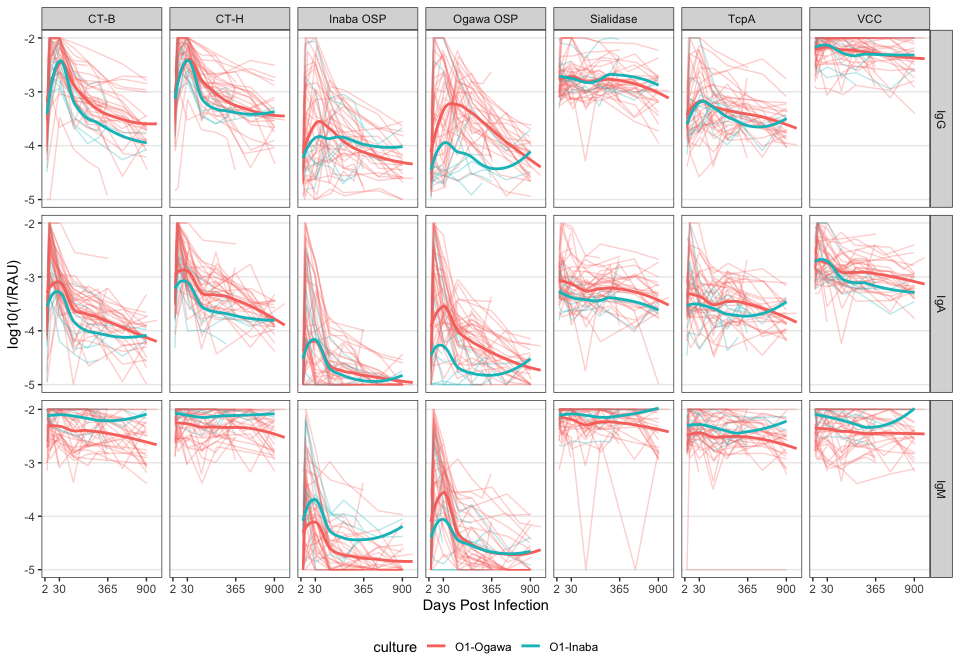

#### Table S4: Relative parameter values for univariate exponential decay models including covariates for different serological markers

The median relative baseline value, median relative fold-change value, and median difference in days of half-life are reported. 95% Bayesian Credible intervals are shown in parentheses. Reference categories include male, non-O blood group, Inaba, and 10+ years old.

| Marker | Covariate | Ratio of Baseline Values | Ratio of Fold-Change Values | Difference in days of Half-Life |
| --- | --- | --- | --- | --- |
| ELISA IgA CtxB | Female | 0.716 (0.473-1.09) | 1.1 (0.509-2.52) | -3 (-13-7) |
|  | O Blood | 0.862 (0.575-1.3) | 1.78 (0.851-3.69) | 9 (1-20)* |
|  | Ogawa | 1.11 (0.645-1.9) | 1.02 (0.382-2.56) | 4 (-11-14) |
|  | Under 10 | 0.809 (0.534-1.23) | 1.93 (0.628-4.05) | 1 (-8-11) |
| ELISA IgA LPS | Female | 1.07 (0.707-1.61) | 0.672 (0.274-1.82) | -7 (-28-35) |
|  | O Blood | 1.31 (0.87-1.96) | 2.12 (0.87-5.02) | -13 (-55-14) |
|  | Ogawa | 1.27 (0.779-2.09) | 1.14 (0.278-3.48) | 18 (-8-39) |
|  | Under 10 | 0.476 (0.337-0.673)* | 0.801 (0.329-1.96) | 12 (-14-57) |
| ELISA IgG CtxB | Female | 0.98 (0.739-1.32) | 1.38 (0.848-3.53) | -30 (-78-5) |
|  | O Blood | 1.05 (0.781-1.4) | 1.34 (0.815-2.48) | -19 (-68-18) |
|  | Ogawa | 0.894 (0.622-1.29) | 0.546 (0.145-1.16) | 36 (-1-78) |
|  | Under 10 | 1.16 (0.872-1.56) | 1.16 (0.7-1.96) | -3 (-49-35) |
| ELISA IgG LPS | Female | 0.892 (0.681-1.17) | 0.922 (0.521-1.56) | 17 (-26-88) |
|  | O Blood | 0.977 (0.744-1.29) | 1.69 (1.04-3.12)* | -28 (-164-23) |
|  | Ogawa | 0.948 (0.677-1.35) | 0.951 (0.22-1.96) | 19 (-55-58) |
|  | Under 10 | 0.793 (0.6-1.05) | 0.684 (0.372-1.17) | 44 (-22-187) |
| RAU IgA CTHT | Female | 1.1 (0.717-1.69) | 0.716 (0.245-2.28) | 1 (-10-18) |
|  | O Blood | 1.11 (0.746-1.67) | 0.708 (0.232-2.05) | 9 (-1-23) |
|  | Ogawa | 1.33 (0.798-2.27) | 1.85 (0.526-6.36) | 9 (-3-20) |
|  | Under 10 | 0.928 (0.611-1.43) | 1.39 (0.45-3.92) | -2 (-14-9) |
| RAU IgA CtxB | Female | 1.18 (0.72-1.91) | 0.61 (0.252-1.42) | 12 (-6-41) |
|  | O Blood | 1.13 (0.684-1.76) | 0.944 (0.411-2.06) | 3 (-11-19) |
|  | Ogawa | 1.26 (0.684-2.34) | 1.99 (0.734-5.37) | 12 (-5-25) |
|  | Under 10 | 0.913 (0.566-1.49) | 1.39 (0.598-3.2) | -6 (-21-9) |
| RAU IgA Flu | Female | 2.81 (0.804-11) | 1.22 (0.389-5.19) | 2 (-9-70) |
|  | O Blood | 1.17 (0.321-4.4) | 0.803 (0.216-2.74) | 0 (-24-15) |
|  | Ogawa | 2.15 (0.472-11.4) | 1.42 (0.348-5.91) | 0 (-85-14) |
|  | Under 10 | 0.343 (0.0928-1.2) | 1.31 (0.37-5.56) | -1 (-37-11) |
| RAU IgA Inaba OSP | Female | 2.41 (1.1-5.72)* | 0.585 (0.147-2.36) | -12 (-27-2) |
|  | O Blood | 1.47 (0.657-3.45) | 3.28 (0.837-12.4) | 9 (-7-23) |
|  | Ogawa | 1.35 (0.455-4.07) | 1.73 (0.38-8.63) | 6 (-15-21) |
|  | Under 10 | 0.306 (0.139-0.618)* | 0.379 (0.104-1.46) | -6 (-20-14) |
| RAU IgA LTB | Female | 0.802 (0.537-1.22) | 1.1 (0.359-4.21) | -2 (-10-10) |
|  | O Blood | 1.31 (0.886-1.92) | 0.709 (0.216-2.15) | 5 (-3-15) |
|  | Ogawa | 1.29 (0.778-2.12) | 1.93 (0.523-7.73) | 1 (-13-11) |
|  | Under 10 | 0.94 (0.633-1.4) | 1.34 (0.41-3.87) | 1 (-8-10) |
| RAU IgA LTh | Female | 1.07 (0.666-1.77) | 0.725 (0.302-1.87) | 3 (-8-20) |
|  | O Blood | 1.37 (0.875-2.17) | 0.669 (0.28-1.58) | 6 (-3-17) |
|  | Ogawa | 1.46 (0.819-2.61) | 1.6 (0.601-4.33) | 2 (-11-12) |
|  | Under 10 | 0.86 (0.541-1.34) | 1.62 (0.669-3.59) | 1 (-9-10) |
| RAU IgA O139 | Female | 1.07 (0.668-1.75) | 0.7 (0.169-2.87) | -3 (-9-5) |
|  | O Blood | 1.18 (0.749-1.86) | 0.556 (0.133-2.25) | 0 (-6-9) |
|  | Ogawa | 1.19 (0.673-2.12) | 1.8 (0.392-7.89) | 0 (-23-6) |
|  | Under 10 | 0.473 (0.322-0.698)* | 1.15 (0.303-4.56) | -2 (-9-4) |
| RAU IgA Ogawa OSP | Female | 1.43 (0.771-2.62) | 0.641 (0.2-2.17) | -12 (-35-17) |
|  | O Blood | 1.33 (0.74-2.42) | 2.57 (0.822-7.82) | -15 (-45-10) |
|  | Ogawa | 2.9 (1.49-5.66)* | 2.82 (0.689-10.3) | 21 (-7-44) |
|  | Under 10 | 0.375 (0.212-0.644)* | 0.402 (0.127-1.2) | 30 (-7-75) |
| RAU IgA Sialidase | Female | 0.968 (0.635-1.49) | 1.08 (0.325-4.13) | -2 (-12-6) |
|  | O Blood | 1.19 (0.797-1.81) | 0.871 (0.252-3.09) | 4 (-4-19) |
|  | Ogawa | 1.47 (0.876-2.47) | 1.42 (0.365-5.14) | 2 (-10-10) |
|  | Under 10 | 0.645 (0.439-0.942)* | 1.55 (0.479-5.79) | -7 (-19-1) |
| RAU IgA TcpA | Female | 1.12 (0.717-1.74) | 1.4 (0.357-6.3) | -5 (-13-2) |
|  | O Blood | 1.03 (0.672-1.58) | 0.61 (0.155-2.34) | 10 (4-21)* |
|  | Ogawa | 1.38 (0.814-2.39) | 0.862 (0.169-4.37) | 5 (-3-13) |
|  | Under 10 | 0.479 (0.34-0.677)* | 0.949 (0.232-4.2) | -8 (-15--2)* |
| RAU IgA VCC | Female | 1.21 (0.796-1.82) | 1.21 (0.285-4.96) | 6 (-1-16) |
|  | O Blood | 1.03 (0.696-1.54) | 0.696 (0.157-2.77) | 5 (-2-15) |
|  | Ogawa | 1.06 (0.63-1.76) | 0.717 (0.136-3.76) | 2 (-9-10) |
|  | Under 10 | 0.617 (0.428-0.884)* | 1.33 (0.321-6.14) | -4 (-12-2) |
| RAU IgG CTHT | Female | 0.559 (0.339-0.925)* | 1.47 (0.607-3.35) | 17 (-22-56) |
|  | O Blood | 0.985 (0.59-1.63) | 0.975 (0.427-2.36) | -13 (-50-25) |
|  | Ogawa | 0.88 (0.459-1.69) | 1.62 (0.493-4.31) | 48 (11-80)* |
|  | Under 10 | 1.91 (1.19-3.11)* | 1.26 (0.541-2.98) | -27 (-71-9) |
| RAU IgG CtxB | Female | 0.848 (0.457-1.54) | 0.974 (0.45-2.03) | 19 (-17-60) |
|  | O Blood | 0.694 (0.383-1.25) | 1.12 (0.543-2.33) | 6 (-30-45) |
|  | Ogawa | 1.26 (0.586-2.71) | 1.4 (0.573-3.5) | 33 (-10-69) |
|  | Under 10 | 1.66 (0.93-3) | 0.951 (0.462-1.96) | -13 (-50-25) |
| RAU IgG Flu | Female | 1.45 (0.313-7.81) | 1.25 (0.479-4.36) | 2 (-28-84) |
|  | O Blood | 1.27 (0.256-6.17) | 0.925 (0.321-2.63) | -1 (-52-153) |
|  | Ogawa | 5.63 (0.859-36.2) | 1.06 (0.273-3.19) | -2 (-138-37) |
|  | Under 10 | 0.351 (0.0744-1.56) | 1.15 (0.451-3.65) | -1 (-113-28) |
| RAU IgG Inaba OSP | Female | 0.904 (0.542-1.5) | 0.921 (0.296-2.84) | -24 (-58-13) |
|  | O Blood | 1.29 (0.778-2.15) | 2.36 (0.812-6.75) | 0 (-47-37) |
|  | Ogawa | 0.99 (0.519-1.88) | 1.12 (0.308-4.01) | 9 (-52-49) |
|  | Under 10 | 0.879 (0.527-1.45) | 0.297 (0.108-0.859)* | 64 (-11-176) |
| RAU IgG LTB | Female | 0.606 (0.371-0.989)* | 1.3 (0.519-2.98) | 13 (-12-41) |
|  | O Blood | 0.875 (0.534-1.44) | 1.02 (0.428-2.57) | 1 (-24-27) |
|  | Ogawa | 1.31 (0.712-2.46) | 1.06 (0.247-3.32) | 29 (3-52)* |
|  | Under 10 | 1.21 (0.75-1.99) | 1.42 (0.601-3.44) | -8 (-35-16) |
| RAU IgG LTh | Female | 0.747 (0.406-1.38) | 1.02 (0.386-2.48) | 17 (-7-44) |
|  | O Blood | 0.993 (0.548-1.79) | 1.03 (0.429-2.58) | 1 (-23-24) |
|  | Ogawa | 1.22 (0.58-2.61) | 1.58 (0.479-4.65) | 26 (0-48)* |
|  | Under 10 | 1.5 (0.835-2.67) | 1.6 (0.662-3.99) | -10 (-33-14) |
| RAU IgG O139 | Female | 0.583 (0.38-0.884)* | 0.9 (0.323-2.99) | -1 (-24-152) |
|  | O Blood | 1.04 (0.67-1.61) | 0.979 (0.395-2.44) | -44 (-154--1)* |
|  | Ogawa | 1.15 (0.664-2.01) | 0.908 (0.216-3.17) | 0 (-42-60) |
|  | Under 10 | 0.79 (0.504-1.21) | 1.47 (0.458-5.08) | -5 (-132-6) |
| RAU IgG Ogawa OSP | Female | 0.636 (0.395-1.05) | 0.947 (0.304-2.98) | 8 (-61-100) |
|  | O Blood | 1.11 (0.672-1.84) | 2.04 (0.656-6.18) | -11 (-90-60) |
|  | Ogawa | 1.94 (1.14-3.37)* | 3.57 (0.994-12.8) | 53 (-16-114) |
|  | Under 10 | 0.894 (0.57-1.42) | 0.281 (0.0955-0.873)* | 175 (71-320)* |
| RAU IgG Sialidase | Female | 0.923 (0.598-1.41) | 1.03 (0.361-3.18) | -1 (-28-119) |
|  | O Blood | 1.17 (0.774-1.76) | 1.06 (0.402-2.59) | -4 (-56-10) |
|  | Ogawa | 0.837 (0.501-1.46) | 1.5 (0.484-5.4) | 1 (-16-51) |
|  | Under 10 | 0.739 (0.501-1.1) | 0.933 (0.244-2.33) | -11 (-90-11) |
| RAU IgG TcpA | Female | 1.14 (0.736-1.78) | 1.17 (0.519-2.73) | -22 (-63-25) |
|  | O Blood | 0.787 (0.512-1.2) | 1.37 (0.616-3.01) | -18 (-89-27) |
|  | Ogawa | 1.35 (0.8-2.28) | 0.632 (0.235-1.66) | 9 (-43-54) |
|  | Under 10 | 0.752 (0.492-1.15) | 0.445 (0.211-1.01) | 27 (-35-129) |
| RAU IgG VCC | Female | 0.915 (0.553-1.51) | 1.42 (0.384-5.41) | 22 (-6-78) |
|  | O Blood | 0.769 (0.476-1.24) | 0.773 (0.134-2.51) | -15 (-90-11) |
|  | Ogawa | 0.938 (0.493-1.72) | 0.942 (0.197-4.12) | -3 (-35-53) |
|  | Under 10 | 0.718 (0.444-1.14) | 1.43 (0.422-6.18) | -4 (-57-17) |
| RAU IgM CTHT | Female | 1.43 (0.87-2.37) | 1.02 (0.257-4.39) | -2 (-15-17) |
|  | O Blood | 0.717 (0.433-1.17) | 0.658 (0.156-2.41) | -2 (-18-9) |
|  | Ogawa | 0.502 (0.276-0.909)* | 1.08 (0.193-5.16) | 2 (-24-15) |
|  | Under 10 | 0.971 (0.565-1.84) | 1.86 (0.423-9.01) | -4 (-69-6) |
| RAU IgM CtxB | Female | 1.45 (0.763-2.74) | 0.718 (0.183-3.2) | 3 (-14-105) |
|  | O Blood | 0.589 (0.318-1.06) | 0.66 (0.141-2.77) | 1 (-24-33) |
|  | Ogawa | 0.411 (0.185-0.881)* | 1.92 (0.409-8.16) | -21 (-194-7) |
|  | Under 10 | 1.02 (0.532-1.91) | 2.81 (0.535-14.5) | -8 (-152-11) |
| RAU IgM Flu | Female | 1.7 (0.938-3.08) | 0.898 (0.26-3.92) | 17 (-10-121) |
|  | O Blood | 0.595 (0.329-1.06) | 0.836 (0.211-2.81) | -1 (-74-23) |
|  | Ogawa | 0.555 (0.253-1.18) | 1.35 (0.331-5.76) | -10 (-140-10) |
|  | Under 10 | 1.26 (0.685-2.29) | 1.25 (0.321-6.1) | 1 (-54-30) |
| RAU IgM Inaba OSP | Female | 2.64 (1.07-6.64)* | 0.762 (0.23-2.65) | -13 (-28-2) |
|  | O Blood | 0.894 (0.331-2.35) | 1.97 (0.629-6.38) | 14 (0-29)* |
|  | Ogawa | 0.285 (0.0878-0.871)* | 1.73 (0.457-6.5) | 12 (-6-27) |
|  | Under 10 | 0.608 (0.237-1.55) | 0.649 (0.198-2.23) | -17 (-30--3)* |
| RAU IgM LTB | Female | 1.37 (0.672-2.85) | 0.939 (0.285-4.06) | 8 (-5-113) |
|  | O Blood | 0.672 (0.333-1.32) | 0.882 (0.231-3.25) | 1 (-27-17) |
|  | Ogawa | 0.449 (0.175-1.05) | 1.01 (0.176-4) | -4 (-87-8) |
|  | Under 10 | 1.18 (0.592-2.38) | 1.21 (0.331-4.9) | -2 (-43-10) |
| RAU IgM LTh | Female | 1.18 (0.604-2.25) | 1.03 (0.283-6.26) | 4 (-6-149) |
|  | O Blood | 0.659 (0.356-1.23) | 0.864 (0.137-3.68) | 1 (-40-17) |
|  | Ogawa | 0.523 (0.239-1.15) | 1.48 (0.287-6.64) | -2 (-177-7) |
|  | Under 10 | 1.1 (0.584-2.04) | 1.27 (0.32-6.22) | 0 (-95-11) |
| RAU IgM O139 | Female | 1.22 (0.685-2.16) | 0.824 (0.351-1.64) | 69 (-5-261) |
|  | O Blood | 0.822 (0.466-1.46) | 1.08 (0.435-2.25) | -38 (-261-23) |
|  | Ogawa | 0.541 (0.263-1.09) | 1.24 (0.471-2.79) | -5 (-347-74) |
|  | Under 10 | 0.563 (0.323-0.973)* | 1.06 (0.471-2.5) | -16 (-233-31) |
| RAU IgM Ogawa OSP | Female | 1.48 (0.605-3.59) | 0.519 (0.17-1.61) | -3 (-29-30) |
|  | O Blood | 1.49 (0.628-3.61) | 1.19 (0.398-3.69) | -11 (-36-14) |
|  | Ogawa | 0.926 (0.303-2.87) | 7.24 (2.02-23.7)* | 25 (-19-48) |
|  | Under 10 | 0.539 (0.23-1.27) | 0.907 (0.295-2.83) | -8 (-32-18) |
| RAU IgM Sialidase | Female | 2 (1.01-3.95)* | 1.08 (0.252-5.51) | 12 (-9-118) |
|  | O Blood | 1.06 (0.519-2.1) | 0.83 (0.182-3.89) | -4 (-64-14) |
|  | Ogawa | 0.534 (0.211-1.32) | 1.51 (0.275-7.76) | -3 (-203-13) |
|  | Under 10 | 0.777 (0.378-1.53) | 1.02 (0.235-5.06) | -1 (-22-26) |
| RAU IgM TcpA | Female | 1.29 (0.47-3.43) | 0.573 (0.17-2.31) | 633 (-8-2154) |
|  | O Blood | 0.864 (0.358-2.09) | 0.936 (0.229-2.94) | -2 (-313-18) |
|  | Ogawa | 0.656 (0.223-1.93) | 1.32 (0.334-4.46) | 2 (-84-24) |
|  | Under 10 | 0.939 (0.396-2.21) | 0.888 (0.273-3.2) | -10 (-295-8) |
| RAU IgM VCC | Female | 1.66 (0.861-3.33) | 1.23 (0.414-4.75) | 4 (-11-45) |
|  | O Blood | 0.558 (0.25-1.08) | 0.784 (0.259-2.28) | -1 (-26-16) |
|  | Ogawa | 0.489 (0.207-1.09) | 0.856 (0.183-2.84) | -3 (-34-14) |
|  | Under 10 | 0.847 (0.425-1.66) | 1.12 (0.383-3.77) | -1 (-30-15) |
| Vibriocidal Inaba | Female | 3.47 (1.12-11.2)* | 0.718 (0.216-2.33) | -33 (-85-27) |
|  | O Blood | 1.35 (0.42-3.94) | 0.839 (0.266-2.53) | 16 (-42-81) |
|  | Ogawa | 0.238 (0.0478-0.962)* | 0.814 (0.212-3.77) | 25 (-37-81) |
|  | Under 10 | 0.901 (0.282-2.84) | 0.587 (0.193-1.84) | 18 (-40-90) |
| Vibriocidal Ogawa | Female | 0.476 (0.173-1.34) | 0.673 (0.176-2.41) | 422 (210-764)* |
|  | O Blood | 1.14 (0.417-3.37) | 1.11 (0.329-3.96) | -36 (-217-82) |
|  | Ogawa | 0.316 (0.0941-1.05) | 3.41 (0.813-13.8) | 102 (-4-244) |
|  | Under 10 | 0.202 (0.0708-0.547)* | 1.48 (0.401-5.45) | 230 (104-439)* |
| IgA NetMFI_CTHT | Female | 0.984 (0.615-1.57) | 0.822 (0.342-2.05) | 0 (-13-21) |
|  | O Blood | 1.08 (0.695-1.69) | 0.839 (0.356-1.92) | 8 (-5-25) |
|  | Ogawa | 1.28 (0.722-2.26) | 1.53 (0.533-4.17) | 12 (-2-25) |
|  | Under 10 | 0.975 (0.629-1.54) | 1.44 (0.632-3.34) | -5 (-20-8) |
| IgA NetMFI_CtxB | Female | 1.08 (0.655-1.76) | 0.695 (0.325-1.47) | 11 (-7-39) |
|  | O Blood | 1.08 (0.662-1.75) | 0.997 (0.471-2.05) | 3 (-11-19) |
|  | Ogawa | 1.14 (0.627-2.06) | 1.97 (0.791-4.78) | 13 (-4-26) |
|  | Under 10 | 0.964 (0.598-1.55) | 1.36 (0.658-2.79) | -6 (-21-9) |
| IgA NetMFI_Flu | Female | 1.83 (0.832-4.06) | 1.18 (0.545-3.9) | 0 (-22-37) |
|  | O Blood | 1.02 (0.469-2.28) | 0.951 (0.359-2.36) | 0 (-36-22) |
|  | Ogawa | 1.83 (0.72-4.89) | 0.974 (0.321-2.42) | -1 (-80-14) |
|  | Under 10 | 0.601 (0.281-1.29) | 1.19 (0.531-3.89) | -1 (-31-18) |
| IgA NetMFI_Inaba OSP | Female | 1.33 (0.724-2.47) | 0.662 (0.161-2.65) | -6 (-18-7) |
|  | O Blood | 1.57 (0.885-2.73) | 2.82 (0.668-9.53) | 8 (-4-20) |
|  | Ogawa | 0.926 (0.431-2.04) | 1.89 (0.397-9.67) | 5 (-13-19) |
|  | Under 10 | 0.477 (0.272-0.831)* | 0.32 (0.0867-1.32) | -8 (-20-9) |
| IgA NetMFI_LTB | Female | 0.742 (0.487-1.15) | 1.08 (0.449-3.02) | -2 (-10-13) |
|  | O Blood | 1.29 (0.862-2) | 0.773 (0.33-1.73) | 4 (-5-15) |
|  | Ogawa | 1.18 (0.689-2.01) | 1.66 (0.58-4.37) | 2 (-12-13) |
|  | Under 10 | 0.935 (0.611-1.44) | 1.45 (0.586-3.33) | 0 (-10-9) |
| IgA NetMFI_LTh | Female | 0.969 (0.619-1.52) | 0.738 (0.349-1.63) | 5 (-6-22) |
|  | O Blood | 1.37 (0.892-2.09) | 0.78 (0.368-1.62) | 6 (-4-16) |
|  | Ogawa | 1.23 (0.707-2.13) | 1.74 (0.708-4.2) | 3 (-11-13) |
|  | Under 10 | 0.951 (0.622-1.47) | 1.49 (0.697-3.18) | 0 (-9-10) |
| IgA NetMFI_O139 | Female | 0.825 (0.529-1.29) | 0.84 (0.234-3.25) | -4 (-11-3) |
|  | O Blood | 1.15 (0.746-1.8) | 0.563 (0.144-2.25) | 1 (-6-12) |
|  | Ogawa | 1.12 (0.636-1.96) | 1.51 (0.325-6.25) | 0 (-24-7) |
|  | Under 10 | 0.487 (0.342-0.695)* | 1.11 (0.31-4.15) | -4 (-10-3) |
| IgA NetMFI_Ogawa OSP | Female | 1.11 (0.691-1.78) | 0.685 (0.206-2.33) | -5 (-24-19) |
|  | O Blood | 1.23 (0.769-1.97) | 2.61 (0.804-8.25) | -11 (-36-10) |
|  | Ogawa | 1.97 (1.17-3.31)* | 3.18 (0.795-12.2) | 16 (-10-35) |
|  | Under 10 | 0.638 (0.409-0.99)* | 0.384 (0.118-1.32) | -12 (-29-17) |
| IgA NetMFI_Sialidase | Female | 0.869 (0.561-1.32) | 1.2 (0.337-4.59) | -2 (-11-4) |
|  | O Blood | 1.23 (0.815-1.88) | 0.728 (0.222-2.5) | 6 (-2-18) |
|  | Ogawa | 1.36 (0.816-2.31) | 1.54 (0.374-5.51) | 2 (-7-9) |
|  | Under 10 | 0.595 (0.412-0.869)* | 1.61 (0.478-6.29) | -8 (-16-0) |
| IgA NetMFI_TcpA | Female | 0.885 (0.587-1.34) | 1.4 (0.355-5.86) | -4 (-12-4) |
|  | O Blood | 1.06 (0.711-1.57) | 0.765 (0.191-2.78) | 9 (2-20)* |
|  | Ogawa | 1.35 (0.82-2.2) | 0.786 (0.165-3.87) | 5 (-4-13) |
|  | Under 10 | 0.616 (0.424-0.877)* | 0.913 (0.238-3.74) | -8 (-16--2)* |
| IgA NetMFI_VCC | Female | 1.09 (0.691-1.71) | 1.58 (0.383-5.85) | 4 (-4-12) |
|  | O Blood | 0.985 (0.626-1.54) | 0.559 (0.154-2.05) | 10 (1-24)* |
|  | Ogawa | 0.911 (0.508-1.58) | 0.764 (0.165-3.77) | 1 (-9-10) |
|  | Under 10 | 0.479 (0.333-0.687)* | 1.1 (0.26-5.94) | -3 (-10-5) |
| IgG NetMFI_CTHT | Female | 0.543 (0.313-0.942)* | 1.85 (0.963-3.52) | 7 (-35-50) |
|  | O Blood | 0.934 (0.526-1.66) | 0.916 (0.485-1.78) | -18 (-60-24) |
|  | Ogawa | 0.909 (0.439-1.89) | 1.36 (0.457-3.27) | 62 (25-97)* |
|  | Under 10 | 2.05 (1.19-3.56)* | 0.907 (0.468-1.77) | -23 (-67-18) |
| IgG NetMFI_CtxB | Female | 0.861 (0.425-1.74) | 1.22 (0.703-2.16) | 4 (-44-54) |
|  | O Blood | 0.67 (0.342-1.28) | 1.25 (0.731-2.2) | -3 (-50-46) |
|  | Ogawa | 1.23 (0.539-2.89) | 1.02 (0.478-2.1) | 64 (18-107)* |
|  | Under 10 | 1.82 (0.949-3.49) | 0.817 (0.471-1.41) | -12 (-61-35) |
| IgG NetMFI_Flu | Female | 1.34 (0.379-4.59) | 1.15 (0.649-2.77) | 2 (-74-148) |
|  | O Blood | 1.04 (0.302-3.56) | 1.03 (0.582-2.11) | 0 (-73-308) |
|  | Ogawa | 5.15 (1.16-23.2)* | 0.748 (0.178-1.66) | -3 (-241-55) |
|  | Under 10 | 0.476 (0.139-1.57) | 1.18 (0.665-2.68) | 0 (-181-57) |
| IgG NetMFI_Inaba OSP | Female | 0.809 (0.397-1.67) | 0.947 (0.273-3.39) | -29 (-64-10) |
|  | O Blood | 1.39 (0.683-2.79) | 2.45 (0.721-7.62) | 1 (-50-41) |
|  | Ogawa | 0.96 (0.386-2.4) | 1.22 (0.299-4.93) | 18 (-44-58) |
|  | Under 10 | 0.99 (0.482-2.05) | 0.262 (0.0835-0.848)* | 70 (-14-193) |
| IgG NetMFI_LTB | Female | 0.601 (0.335-1.08) | 1.57 (0.74-3.29) | 10 (-19-40) |
|  | O Blood | 0.829 (0.461-1.49) | 0.929 (0.442-1.92) | 0 (-29-28) |
|  | Ogawa | 1.54 (0.746-3.08) | 0.937 (0.231-2.47) | 35 (9-59)* |
|  | Under 10 | 1.23 (0.687-2.17) | 1.28 (0.629-2.62) | -5 (-34-23) |
| IgG NetMFI_LTh | Female | 0.783 (0.392-1.55) | 1.51 (0.768-2.95) | 11 (-18-43) |
|  | O Blood | 0.946 (0.488-1.85) | 0.855 (0.438-1.67) | -3 (-33-26) |
|  | Ogawa | 1.36 (0.608-3.08) | 1.04 (0.365-2.47) | 40 (11-66)* |
|  | Under 10 | 1.53 (0.789-2.92) | 1.08 (0.557-2.09) | -1 (-31-29) |
| IgG NetMFI_O139 | Female | 0.584 (0.363-0.935)* | 0.969 (0.318-3.36) | -2 (-17-61) |
|  | O Blood | 0.944 (0.569-1.55) | 0.889 (0.219-2.45) | -21 (-130-2) |
|  | Ogawa | 1.34 (0.737-2.46) | 0.792 (0.179-2.83) | 1 (-27-21) |
|  | Under 10 | 0.712 (0.443-1.15) | 1.77 (0.54-6.53) | -3 (-55-7) |
| IgG NetMFI_Ogawa OSP | Female | 0.63 (0.389-1.04) | 1.02 (0.315-3.19) | -14 (-77-63) |
|  | O Blood | 1.04 (0.621-1.75) | 1.98 (0.62-5.86) | 4 (-73-68) |
|  | Ogawa | 1.92 (1.1-3.48)* | 3.97 (1.11-14.3)* | 51 (-15-107) |
|  | Under 10 | 0.909 (0.566-1.48) | 0.315 (0.104-0.969)* | 142 (45-285)* |
| IgG NetMFI_Sialidase | Female | 0.87 (0.557-1.36) | 1.27 (0.588-3.34) | -17 (-37-144) |
|  | O Blood | 1.1 (0.722-1.71) | 0.928 (0.501-1.86) | -6 (-39-32) |
|  | Ogawa | 0.853 (0.503-1.46) | 0.976 (0.371-2.09) | 27 (10-51)* |
|  | Under 10 | 0.762 (0.502-1.15) | 1.07 (0.533-2.62) | -36 (-67--16)* |
| IgG NetMFI_TcpA | Female | 0.902 (0.542-1.53) | 1.08 (0.46-2.56) | -21 (-55-19) |
|  | O Blood | 0.784 (0.471-1.29) | 1.41 (0.613-3.12) | -11 (-72-27) |
|  | Ogawa | 1.49 (0.804-2.79) | 0.565 (0.206-1.5) | 5 (-36-44) |
|  | Under 10 | 1.1 (0.669-1.8) | 0.42 (0.194-0.959)* | 8 (-38-93) |
| IgG NetMFI_VCC | Female | 0.765 (0.47-1.22) | 1.15 (0.312-2.68) | 30 (-15-82) |
|  | O Blood | 0.749 (0.474-1.18) | 0.915 (0.452-2.11) | -19 (-70-26) |
|  | Ogawa | 0.973 (0.529-1.76) | 0.594 (0.121-1.4) | 33 (-9-86) |
|  | Under 10 | 0.909 (0.579-1.43) | 1.64 (0.687-5.01) | -38 (-70--11)* |
| IgM NetMFI_CTHT | Female | 1.48 (1-2.2) | 1.28 (0.427-5.16) | -2 (-15-8) |
|  | O Blood | 0.788 (0.537-1.18) | 0.754 (0.238-2.41) | 2 (-7-20) |
|  | Ogawa | 0.71 (0.444-1.2) | 1.3 (0.371-4.25) | 3 (-16-15) |
|  | Under 10 | 1.13 (0.753-1.75) | 1.28 (0.394-4.52) | -2 (-21-8) |
| IgM NetMFI_CtxB | Female | 1.45 (0.943-2.28) | 1.1 (0.386-5.2) | 0 (-38-37) |
|  | O Blood | 0.655 (0.427-1) | 1.01 (0.32-2.86) | 1 (-95-22) |
|  | Ogawa | 0.663 (0.378-1.14) | 1.58 (0.432-5.87) | -7 (-112-14) |
|  | Under 10 | 1.05 (0.681-1.63) | 1.33 (0.466-4.42) | -2 (-95-13) |
| IgM NetMFI_Flu | Female | 1.49 (0.933-2.38) | 2.27 (0.666-9.42) | 0 (-8-7) |
|  | O Blood | 0.661 (0.416-1.04) | 0.691 (0.222-2.04) | 1 (-4-11) |
|  | Ogawa | 0.841 (0.457-1.54) | 0.387 (0.075-1.79) | 2 (-5-14) |
|  | Under 10 | 1.32 (0.832-2.12) | 1.75 (0.549-6.55) | -2 (-11-3) |
| IgM NetMFI_Inaba OSP | Female | 2.01 (1.08-3.67)* | 0.924 (0.308-2.87) | -9 (-23-5) |
|  | O Blood | 0.895 (0.483-1.71) | 1.91 (0.639-5.35) | 13 (0-26) |
|  | Ogawa | 0.467 (0.214-1.03) | 1.18 (0.341-4.3) | 8 (-9-21) |
|  | Under 10 | 0.806 (0.431-1.48) | 0.532 (0.181-1.7) | -16 (-28--3)* |
| IgM NetMFI_LTB | Female | 1.29 (0.822-1.99) | 1.15 (0.437-4.06) | 1 (-9-81) |
|  | O Blood | 0.807 (0.531-1.24) | 1.1 (0.389-3.09) | 3 (-14-20) |
|  | Ogawa | 0.681 (0.403-1.19) | 1.09 (0.304-3.08) | 0 (-39-12) |
|  | Under 10 | 1.3 (0.85-1.98) | 1.16 (0.44-3.74) | -2 (-44-8) |
| IgM NetMFI_LTh | Female | 1.19 (0.81-1.76) | 1.19 (0.419-4.94) | 2 (-5-20) |
|  | O Blood | 0.744 (0.513-1.09) | 1.04 (0.339-3.23) | 2 (-4-14) |
|  | Ogawa | 0.759 (0.469-1.23) | 1.33 (0.353-4.3) | 0 (-19-7) |
|  | Under 10 | 1.12 (0.76-1.63) | 1.28 (0.44-4.41) | -1 (-13-6) |
| IgM NetMFI_O139 | Female | 1.29 (0.724-2.24) | 0.933 (0.483-1.71) | 68 (-7-277) |
|  | O Blood | 0.866 (0.501-1.49) | 1.07 (0.571-2) | -28 (-262-31) |
|  | Ogawa | 0.575 (0.297-1.11) | 1.16 (0.535-2.25) | -2 (-247-114) |
|  | Under 10 | 0.611 (0.363-1.04) | 1.09 (0.604-2.34) | -8 (-187-58) |
| IgM NetMFI_Ogawa OSP | Female | 1.15 (0.627-2.14) | 0.632 (0.22-1.84) | 5 (-15-32) |
|  | O Blood | 1.31 (0.715-2.37) | 1.29 (0.448-3.8) | -7 (-27-13) |
|  | Ogawa | 1.1 (0.514-2.33) | 5.16 (1.57-16)* | 19 (-15-37) |
|  | Under 10 | 0.927 (0.503-1.67) | 0.696 (0.25-1.95) | -21 (-41--3)* |
| IgM NetMFI_Sialidase | Female | 1.37 (0.943-1.94) | 0.979 (0.552-1.85) | 6 (-21-110) |
|  | O Blood | 0.898 (0.627-1.3) | 1.04 (0.547-1.94) | -1 (-158-25) |
|  | Ogawa | 0.73 (0.469-1.15) | 0.908 (0.366-1.68) | -4 (-87-25) |
|  | Under 10 | 0.909 (0.619-1.32) | 1.06 (0.593-1.99) | 0 (-44-72) |
| IgM NetMFI_TcpA | Female | 1.58 (1.01-2.42)* | 1.36 (0.498-4.76) | -3 (-15-5) |
|  | O Blood | 0.791 (0.511-1.23) | 0.738 (0.249-2.08) | 3 (-5-17) |
|  | Ogawa | 0.837 (0.482-1.48) | 1.1 (0.303-3.15) | 1 (-16-9) |
|  | Under 10 | 0.946 (0.604-1.45) | 0.794 (0.305-2.31) | -2 (-10-8) |
| IgM NetMFI_VCC | Female | 1.85 (1.2-2.84)* | 0.959 (0.548-1.89) | 7 (-9-76) |
|  | O Blood | 0.682 (0.44-1.07) | 1.05 (0.5-2.01) | 2 (-39-28) |
|  | Ogawa | 0.711 (0.397-1.25) | 1.04 (0.401-1.87) | -3 (-63-16) |
|  | Under 10 | 0.909 (0.574-1.45) | 1.09 (0.598-2.02) | 0 (-36-25) |

#### Figure S20: Estimated duration of half-life and average fold-change from exponential decay models fit to Net MFI measurements

Each point indicates the median estimate of the average individual fold-rise from baseline to peak (y-value) and the median estimate of the half-life (x-value) for exponential decay univariate models. Marginal 95% credible intervals are shown as lines. Model estimates for the vibriocidal assay are shown for reference and are identical across panels.

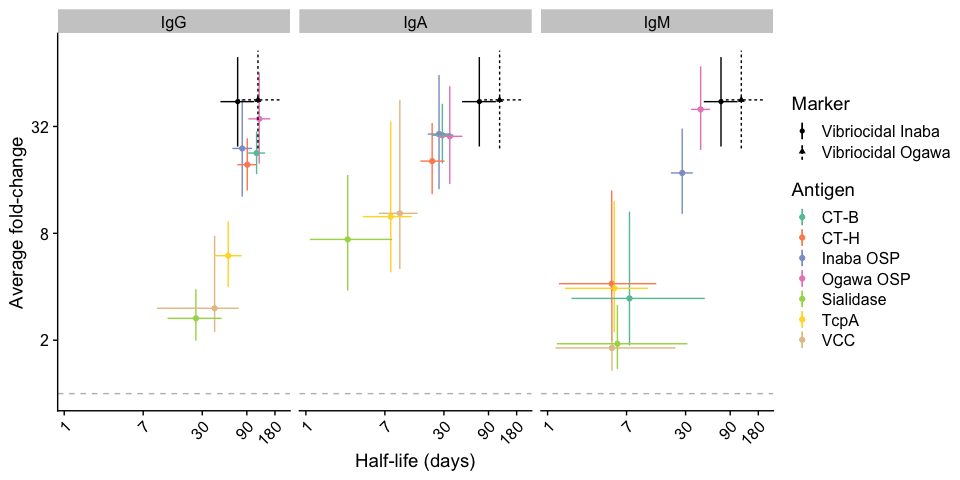

#### Figure S21: Cross-validated receiver operator characteristic curves and permutation importance for random forest models using 45-day, 120-day, 200-day, and 300-day infection window

Permutation importance is shown along the x-axis for 21 predictors of random forest models containing multiplex bead assay (MBA) markers (except for those binding to CT-H, LT-H, and LT-B), age, sex, and blood-type.

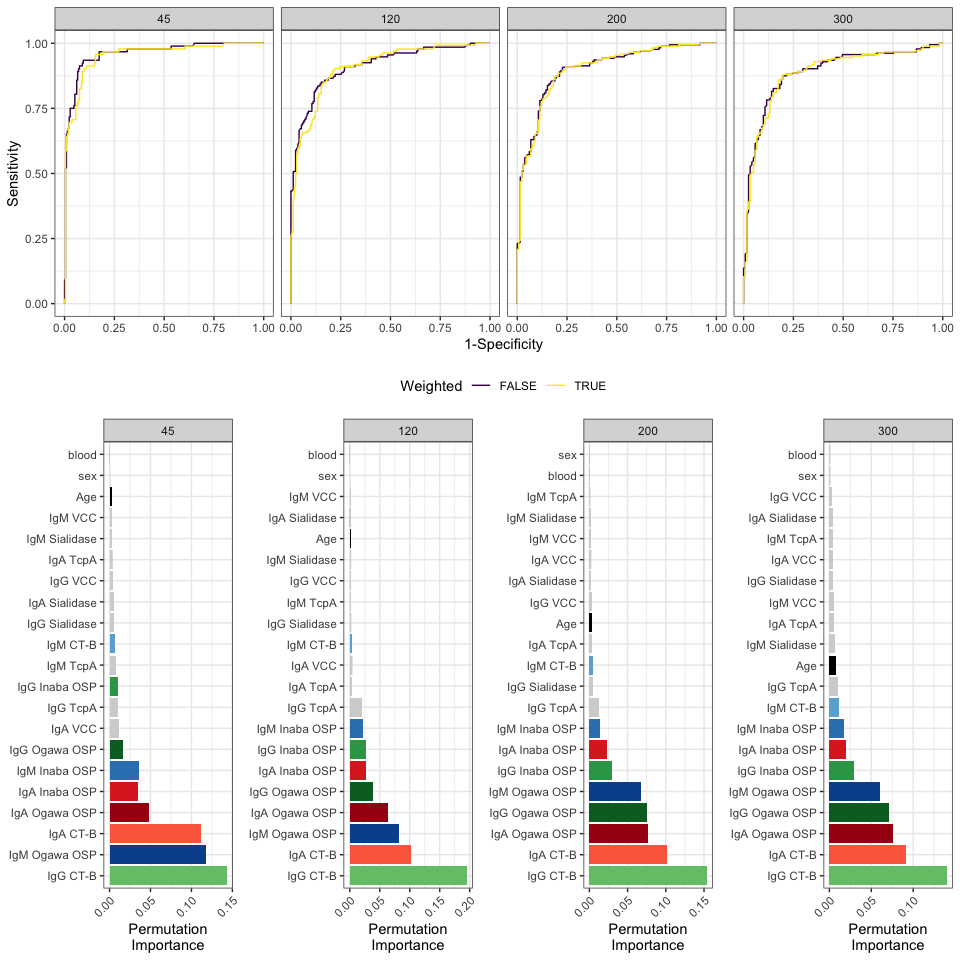

#### Figure S22: Comparison of cross-validated area under the ROC curve between ensemble and random forest models

Models were fit to 18 MBA markers and 3 demographic variables (age, sex, and blood type). The ensemble model was created using the R package SuperLearner. Four different types of models were combined into the ensemble: Random Forest models (ranger), Lasso and Elastic-Net Regularized Generalized Linear Models (glmnet), Bayesian Additive Regression Trees (bartMachine), and Extreme Gradient Boosting (xgboost). All models were unweighted.

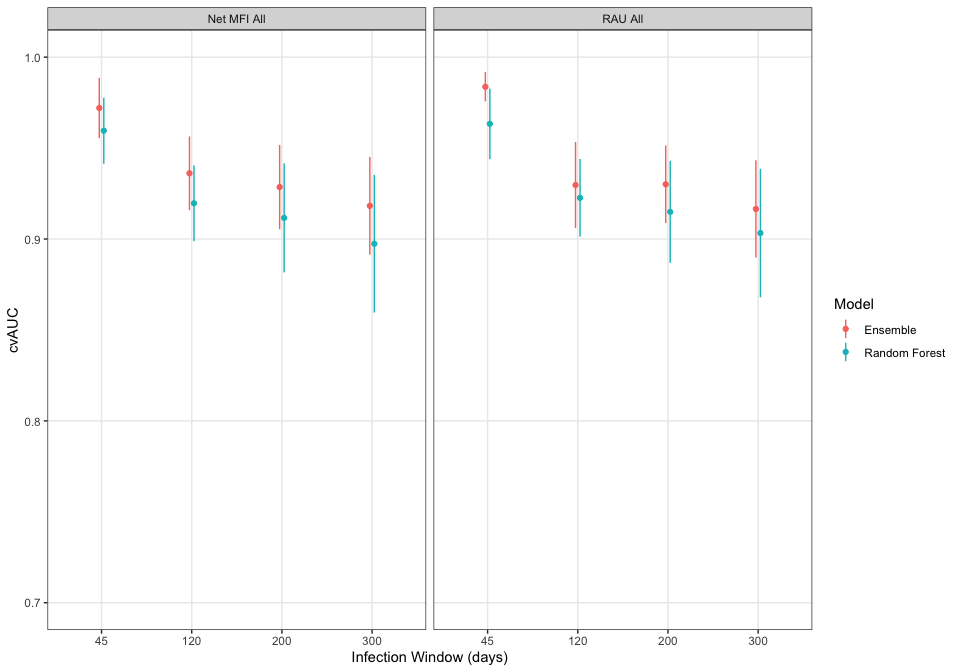

#### Figure S23: Cross-validated area under the receiver operating characteristic curve (cvAUC) and predictor importance rankings for Net MFI markers across random forest models with varying infection windows

Estimates of mean cvAUC (10-fold) and 95% confidence interval are shown for weighted and non-weighted models between 50- and 600-day infection windows at 10-day intervals (A). Rug plot shows the day of collection of samples from cases used in training models. Samples collected under 5 days since infection, over 600 days since infection, or from household contacts are not shown. For each infection window of weighted models, the rankings of predictors by their importance are shown on the y-axis (B). Colors of lines are unique to each predictor.

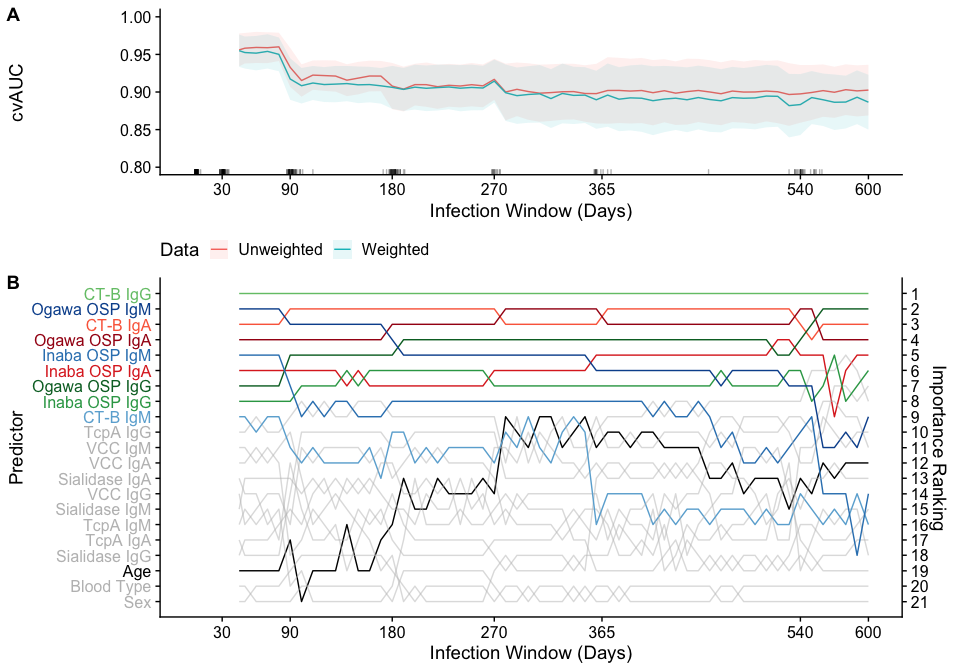

#### Table S5: Comparison of cross-validated AUC between multiple marker random forest models using 45-day, 120-day, 200-day, and 300-day infection windows

Random forest models were fit using a the specified marker set and individual level factors including age, sex, and blood group. Mean and 95% confidence intervals for cvAUC are reported.

| Marker Set | 45-day | 120-day | 200-day | 300-day |
| --- | --- | --- | --- | --- |
| All NetMFI Markers | 0.96 (0.94-0.98) | 0.91 (0.89-0.93) | 0.91 (0.88-0.94) | 0.90 (0.86-0.93) |
| All RAU Markers | 0.95 (0.93-0.97) | 0.92 (0.89-0.94) | 0.92 (0.89-0.94) | 0.91 (0.87-0.94) |
| ELISA Markers | 0.94 (0.91-0.98) | 0.87 (0.84-0.91) | 0.80 (0.76-0.85) | 0.78 (0.74-0.83) |
| IgA NetMFI Markers | 0.92 (0.90-0.95) | 0.89 (0.86-0.92) | 0.89 (0.86-0.92) | 0.86 (0.82-0.91) |
| IgA RAU Markers | 0.93 (0.90-0.96) | 0.89 (0.86-0.91) | 0.89 (0.87-0.92) | 0.84 (0.81-0.87) |
| IgG NetMFI Markers | 0.93 (0.90-0.96) | 0.90 (0.87-0.93) | 0.90 (0.87-0.93) | 0.87 (0.83-0.91) |
| IgG RAU Markers | 0.92 (0.89-0.95) | 0.90 (0.87-0.93) | 0.91 (0.88-0.93) | 0.88 (0.84-0.92) |
| IgM NetMFI Markers | 0.90 (0.86-0.95) | 0.83 (0.80-0.87) | 0.78 (0.73-0.82) | 0.77 (0.72-0.82) |
| IgM RAU Markers | 0.90 (0.86-0.94) | 0.85 (0.81-0.88) | 0.80 (0.76-0.84) | 0.80 (0.75-0.85) |
| Vibriocidal Markers | 0.92 (0.88-0.96) | 0.86 (0.81-0.90) | 0.83 (0.78-0.88) | 0.81 (0.76-0.85) |
| Vibriocidal & ELISA Markers | 0.97 (0.95-0.99) | 0.92 (0.90-0.94) | 0.88 (0.85-0.91) | 0.88 (0.85-0.91) |

#### Table S6: Comparison of cross-validated AUC between multiplex bead assay IgG multiple marker random forest models using 45-day, 120-day, 200-day, and 300-day infection windows

Random forest models were using a reduced panel of MBA IgG markers and individual level factors including age, sex, and blood group. Mean and 95% confidence intervals for cvAUC are reported.

| MBA Panel for IgG | 45-day | 120-day | 200-day | 300-day |
| --- | --- | --- | --- | --- |
| CT-B | 0.89 (0.83-0.95) | 0.86 (0.81-0.90) | 0.82 (0.78-0.86) | 0.77 (0.71-0.83) |
| + Ogawa OSP | 0.91 (0.87-0.95) | 0.89 (0.86-0.92) | 0.88 (0.85-0.91) | 0.84 (0.80-0.88) |
| + Inaba OSP | 0.93 (0.89-0.96) | 0.90 (0.87-0.93) | 0.89 (0.87-0.92) | 0.87 (0.83-0.91) |
| + TcpA | 0.93 (0.90-0.96) | 0.90 (0.88-0.93) | 0.89 (0.87-0.92) | 0.89 (0.85-0.92) |
| + Sialidase | 0.93 (0.90-0.96) | 0.91 (0.88-0.93) | 0.91 (0.89-0.94) | 0.89 (0.86-0.93) |
| + VCC | 0.93 (0.90-0.96) | 0.90 (0.87-0.93) | 0.91 (0.88-0.94) | 0.87 (0.83-0.92) |

#### Figure S24: Comparison of cross-validated AUC across random forest models trained on traditional and Net MFI MBA serological markers for 45-day, 120-day, 200-day, and 300-day infection windows

Random forest models were fit using a specified marker set and individual level factors including age, sex, and blood type (A). Estimated mean and 95% confidence intervals for cvAUC are reported. Models fit to reduced panels of IgG MBA markers are shown (B). The order of how antigens were added was determined by the variable importance when fitting a model with only IgG MBA markers.

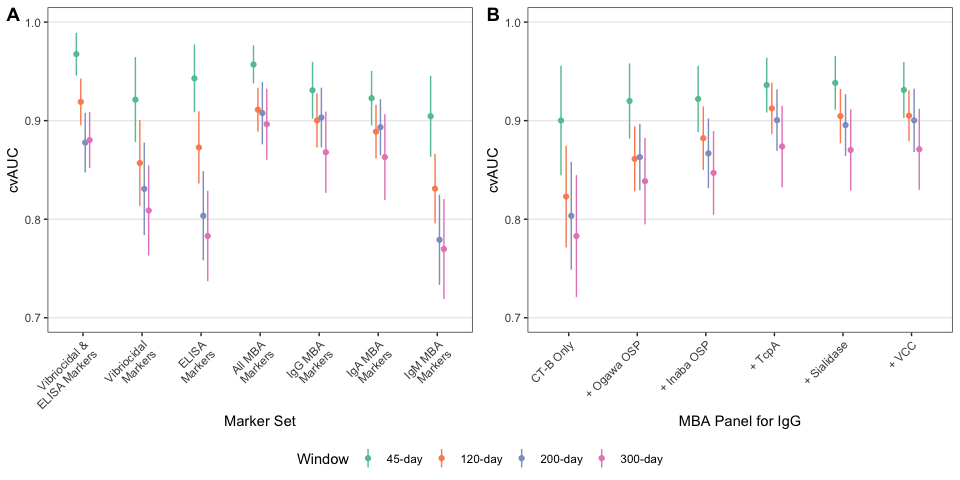

#### Figure S25: Specificity and time-varying sensitivity estimates of random forest models trained on traditional and Net MFI MBA serological markers with leave-one-out cross-validation for 45-day, 120-day, 200-day, and 300-day infection windows using different cut-offs

Median and 95% credible intervals are shown for the estimated (A) nominal specificity (black dashed line) and (B) time-varying sensitivity. Each row represents a different method for acquiring a cut-off including the Youden Index or maximizing sensitivity for a desired value of specificity. The relationship between logit(sensitivity) and time since infection (log-transformed) was constant for the 45-day window, linear for the 120-day, quadratic for the 200-day window, and cubic for the 300-day window. Traditional = vibriocidal Ogawa, vibriocidal Inaba, and 4 ELISA markers, All MBA = 18 MBA markers, All MBA IgG = 6 MBA markers, Reduced panel= Ogawa OSP, Inaba OSP, and CT-B IgG. All models also included age, sex, and blood type as predictors.

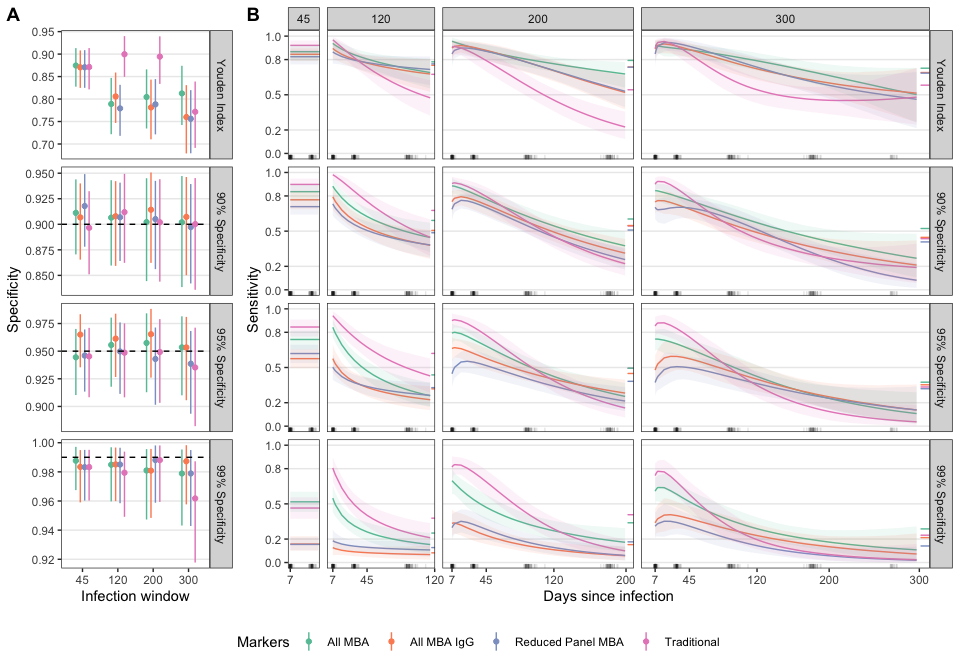

#### Figure S26: Comparison of cross-validated AUC across random forest models trained on traditional and MBA serological markers excluding anti-CT-B markers for 45-day, 120-day, 200-day, and 300-day infection windows

(A) Random forest models were fit using a the specified marker set and individual level factors including age, sex, and blood group. Estimated mean and 95% confidence intervals for cvAUC are reported. (B-D) Models fit to reduced panels of IgG, IgA, and IgM MBA markers are shown.

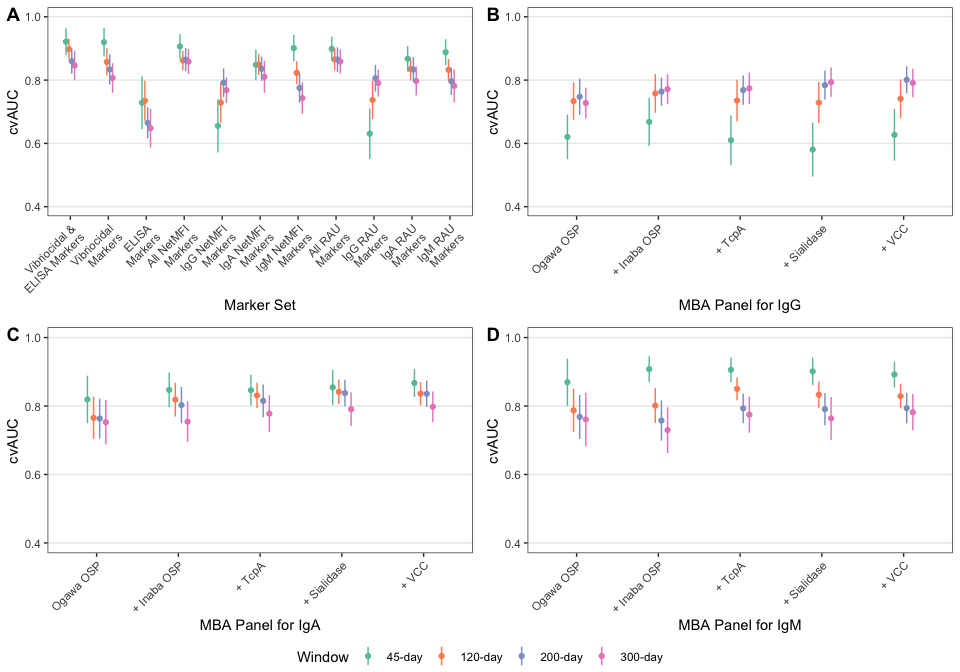
